# Smoking is associated with significantly increased green autofluorescence intensity and asymmetry of the skin and the fingernails of natural populations, population high-risk of developing stroke, and population of acute ischemic stroke

**DOI:** 10.1101/2020.07.24.20161620

**Authors:** Mingchao Zhang, Yue Tao, Danhong Wu, Weihai Ying

## Abstract

Tobacco smoking is an important risk factor for numerous diseases. It is critically needed to search for the biomarkers of smoking for non-invasive and rapid monitoring of the pathological changes of smokers’ body. Our current study has indicated that green autofluorescence (AF) of the fingernails and certain locations of the skin is a novel biomarker for smoking: First, for the natural population at age between 20 - 50 years of old, both the green AF intensity and the AF asymmetry of the Index Fingernails and the skin of Dorsal Index Fingers of the smokers were remarkably higher than those of the non-smokers. Second, for the natural population, the population at risk of developing acute ischemic stroke (AIS) and the AIS population at age between 50 - 80 years of old, both the AF intensity and the AF asymmetry of the Index Fingernails and the skin of Dorsal Index Fingers of the smokers were also remarkably higher than those of the non-smokers. Third, ROC analyses using the green AF intensity of the Index Fingernails showed that the AUC values were 0.796 to 0.889 for differentiating the smokers and the non-smokers in these three populations. Collectively, our study has indicated that increased green AF intensity of the fingernails and certain locations of the skin is a novel biomarker for smoking. Based on this finding, pathological alterations of smokers’ body may be monitored non-invasively and efficiently, which could be highly valuable for the health management of the large population of tobacco smokers.

## Introduction

Tobacco smoking is an important risk factor for multiple major diseases including cancer, AIS and cardiovascular diseases (1-3). In many Western countries smoking is responsible for one third of all cancer deaths (4). It is of significance to search for the biomarker of smoking for non-invasive and rapid monitoring of the pathological changes of smokers’ body, which is highly valuable for the health monitoring of a large population of smokers. However, there has been no non-invasive method for monitoring the pathological changes of smokers.

Human autofluorescence (AF) has been used for non-invasive diagnosis of diabetes and diabetes-related pathology (5). The AF originates from the advanced glycation end-product (AGE)-modified collagen and elastin of dermis. NADH, FAD, keratins and melanin are major epidermal fluorophores (6, 7). It is established that such molecules as NADH and FAD can be significantly affected by oxidative stress.

Since tobacco smoking can induce oxidative stress in the body (8, 9), we hypothesized that tobacco smoking-induced changes of the skin’s AF may become a novel biomarker for tobacco smoking. In this study we tested this hypothesis, showing that smokers had significantly increased green AF intensity and asymmetry of the Index Fingernails and the skin of Dorsal Index Fingers in natural populations, the population high-risk of developing stroke and the population of AIS.

## Methods and Materials

### Studies on Human Subjects

The study was conducted according to a protocol approved by the Ethics Committee of Shanghai Fifth People’s Hospital affiliated to Fudan University. The human subjects of the age group between 20 - 50 years of old came from 郑州新密队列。The age of group was 34.40 ± 7.59 years of old. The human subjects of the age group between 50 - 80 years of old were divided into four groups: Group 1: The natural populations; Group 2: The healthy population; Group 3: The population high-risk of developing stroke; and Group 4: The population of the AIS patients who were hospitalized in the Department of Neurology, Shanghai Fifth People’s Hospital affiliated to Fudan University. The age of Group 1, Group 2, Group 3 and Group 4 was 66.10 ± 6.42, 62.86 ± 5.73, 66.12 ± 5.18, and 64.01±8.20 years of old, respectively.

### Determinations of the AF of human subjects’ skin and fingernails

A portable AF imaging equipment was used to determine the skin’ AF of the human subjects. The excitation wavelength was 485 nm, and the emission wavelength was 500 - 550 nm.

### Statistical analyses

All data are presented as mean ± SEM. Data were assessed by Kruskal-Wallis test, followed by Student - Newman - Keuls *post hoc* test, except where noted. *P* values less than 0.05 were considered statistically significant.

## Results

### 1) Both the green AF intensity and the AF asymmetry of the fingernails and the skin of Dorsal Index Fingers of the smokers in natural populations were significantly higher than those of the non-smokers

We determined the green AF intensity of the fingernails and twelve positions of the skin of natural populations of the age group between 20 - 50 years of old and the age group between 50 - 80 years of old. For both of these age groups, compared with the green AF intensity of the Index Fingernails and the skin of Dorsal Index Fingers of the non-smokers, the green AF intensity of both the Index Fingernails and the skin of Dorsal Index Fingers of the smokers was significantly higher (Figs. 1A and 1B). There was no significant difference between the AF intensity of the left Index Fingernails and skin of Dorsal Index Fingers and that of the right Index Fingernails and skin of Dorsal Index Fingers (Figs. 1A and 1B). In contrast, no significant difference was observed between the green AF intensity of the non-smokers and that of the smokers of these two age groups at other examined positions of the skin (Supplemental Figs. 1A, 1B, 1C, 1D, and 1E).

**Fig. 1.**
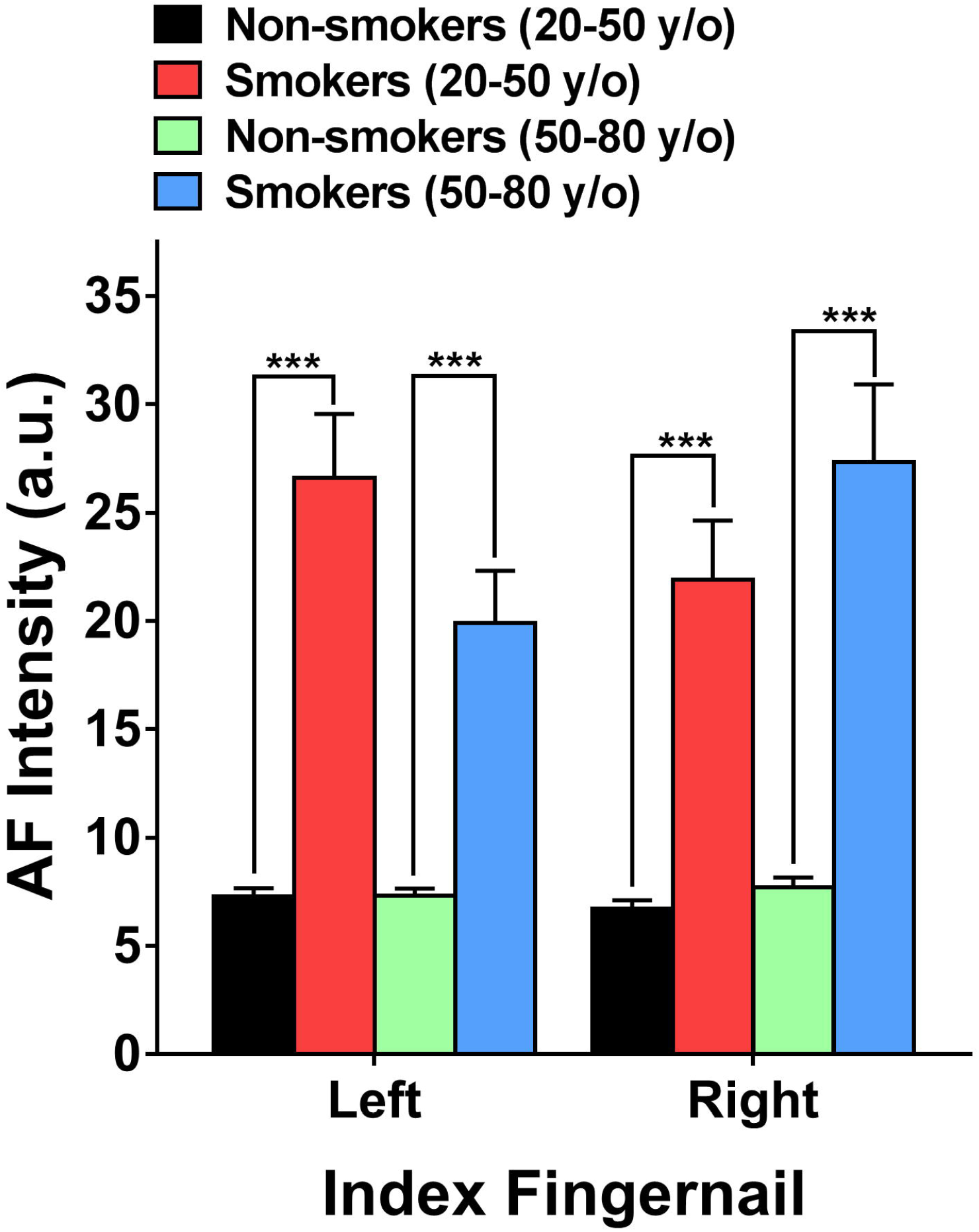
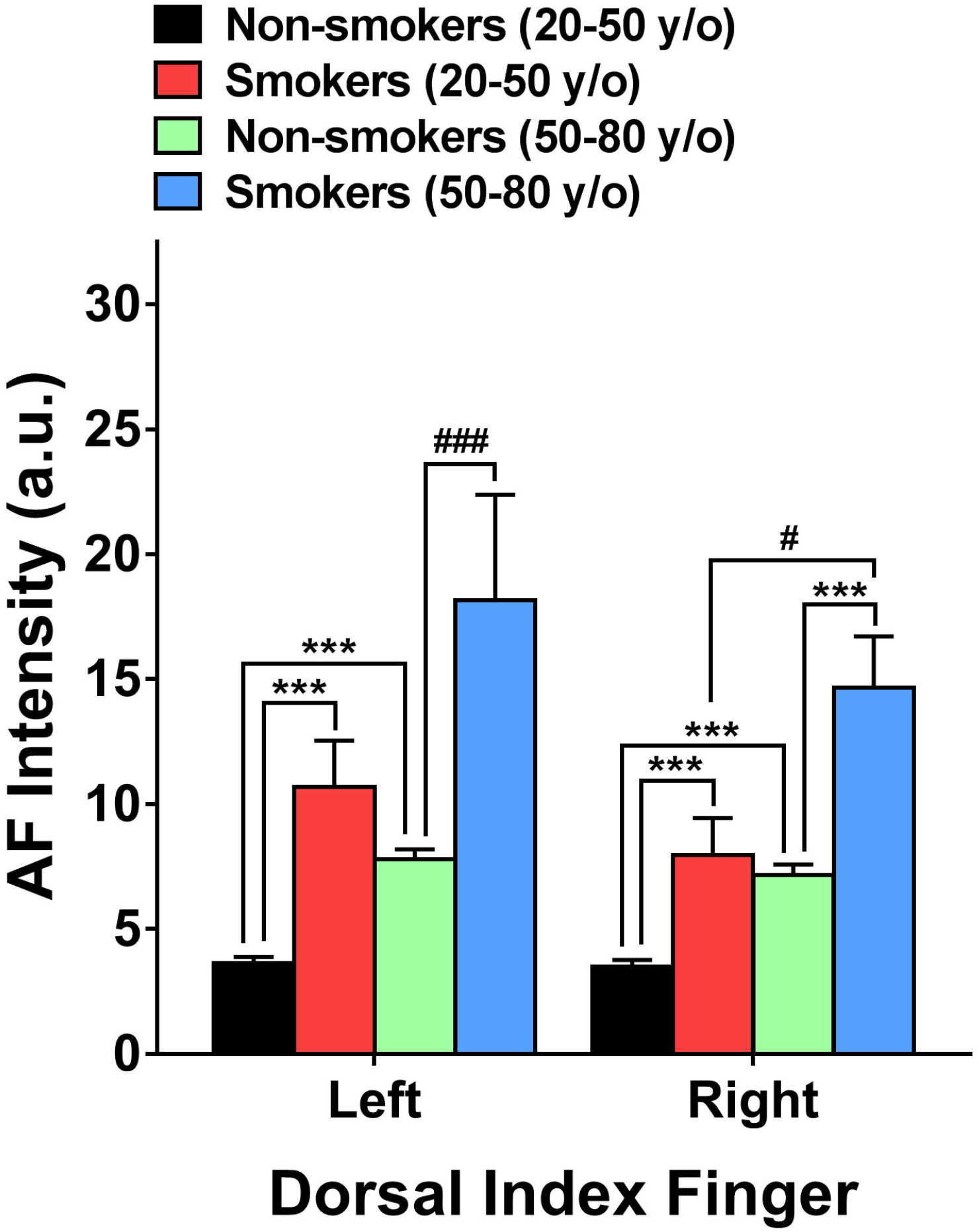
The green AF intensity of the Index Fingernails and the skin of Dorsal Index Fingers of the smokers in natural populations was significantly higher than that of the non-smokers. (A) For both the age group between 20 - 50 years of old and the age group between 50 - 80 years of old, the AF intensity of the smokers in natural populations in their right and left Index Fingernails was significantly higher than that of the non-smokers. (B) For both the age group between 20 - 50 years of old and the age group between 50 - 80 years of old, the AF intensity of the smokers in natural populations in their right and left Dorsal Index Fingers was significantly higher than that of the non-smokers. The number of the non-smokers and the smokers in the age group between 20 - 50 years of old was 165-170 and 23, respectively. The number of the non-smokers and the smokers in the age group between 50 - 80 years of old was 242-246 and 36-38, respectively. ***, *p* < 0.001; #, *p* < 0.05 (Mann-Whitney test); ###, *p* < 0.001 (Mann-Whitney test).

For both of these two age groups, compared with the green AF asymmetry of the fingernails and the skin of Dorsal Index Fingers of the non-smokers, the green AF asymmetry of both the Index Fingernails and the skin of Dorsal Index Fingers of the smokers was significantly higher (Figs. 2A and 2B). In contrast, no significant difference was observed between the green AF asymmetry of the non-smokers and that of the smokers of these two age groups at other examined positions of the skin (Supplemental Figs. 2A, 2B, 2C, 2D, and 2E).

**Fig. 2.**
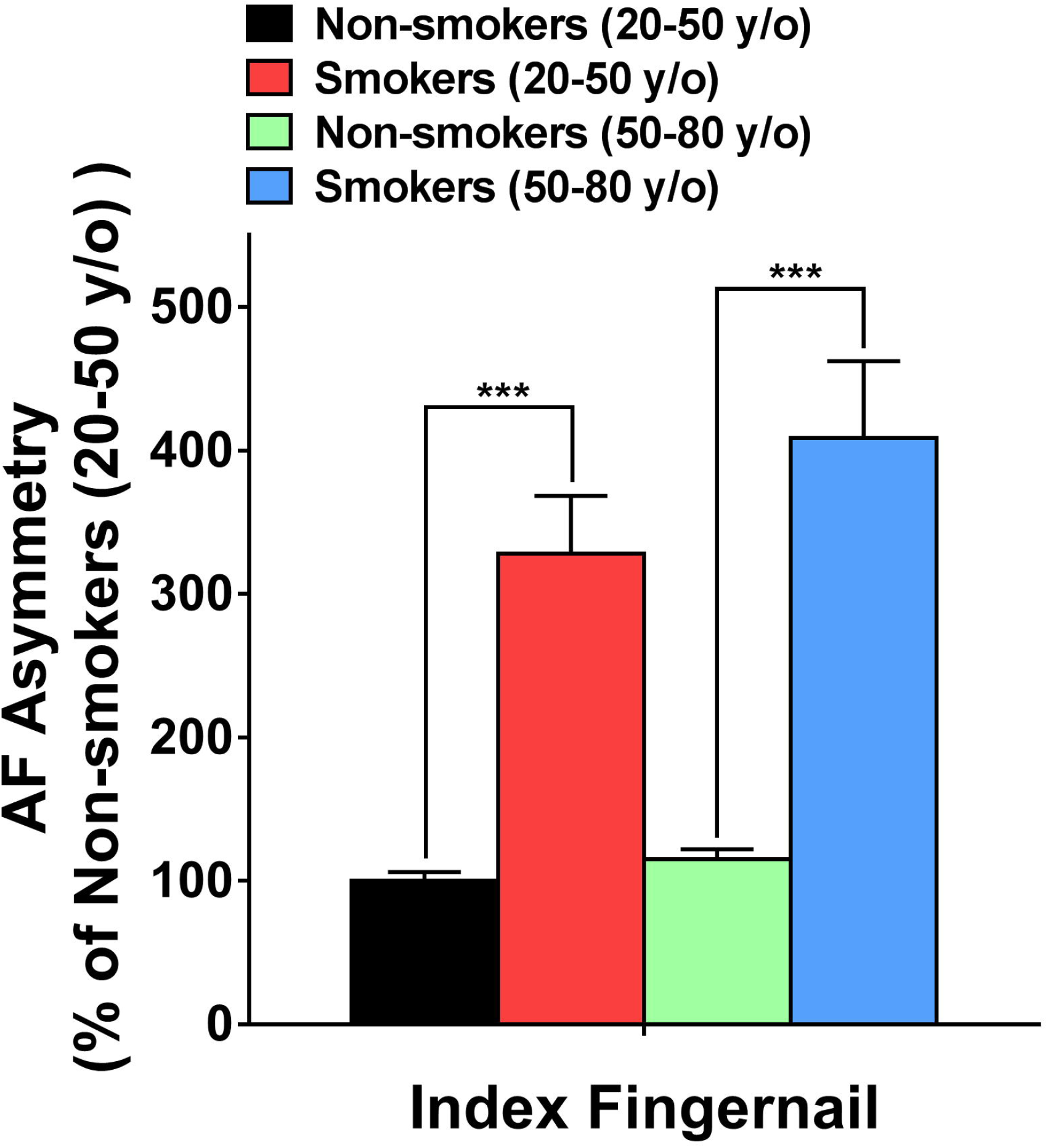
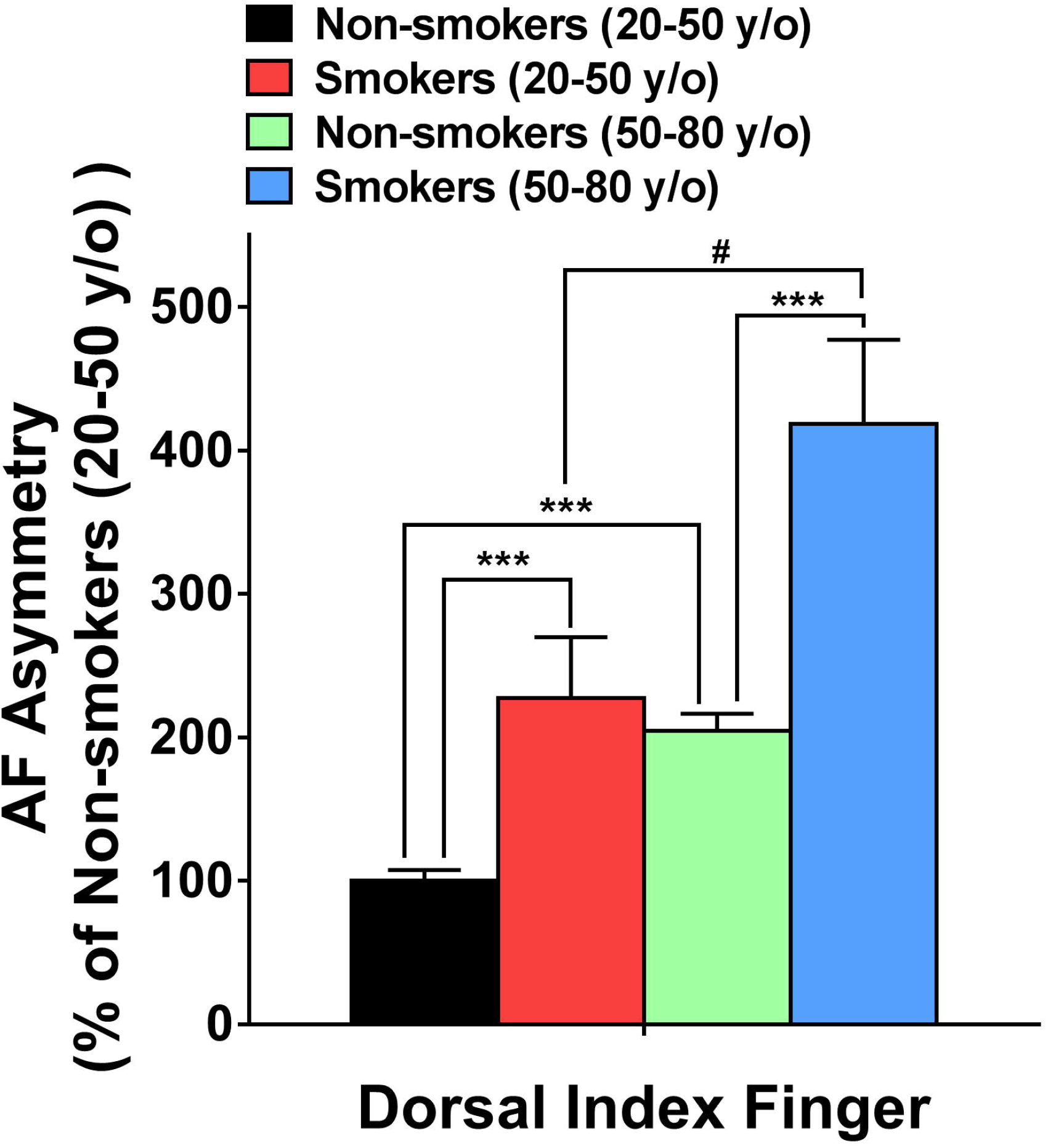
The green AF asymmetry of the Index Fingernails and the skin of Dorsal Index Fingers of the smokers in natural populations was significantly higher than that of the non-smokers. (A) For both the age group between 20 - 50 years of old and the age group between 50 - 80 years of old, the AF asymmetry of the smokers in natural populations in their right and left Index Fingernails was significantly higher than that of the non-smokers. (B) For both the age group between 20 - 50 years of old and the age group between 50 - 80 years of old, the AF asymmetry of the smokers in natural populations in their right and left Dorsal Index Fingers was significantly higher than that of the non-smokers. The number of the non-smokers and the smokers in the age group between 20 - 50 years of old was 165-170 and 23, respectively. The number of the non-smokers and the smokers in the age group between 50 - 80 years of old was 243-246 and 36-38, respectively. ***, *p* < 0.001; #, *p* < 0.05 (Mann-Whitney test).

### 2) The green AF intensity of the Index Fingernails and the skin of Dorsal Index Fingers of the smokers in both the population high-risk of developing stroke and the AIS population was significantly higher than that of the non-smokers

We also determined the green AF intensity of the Index Fingernails and twelve positions of the skin of the population high-risk of developing stroke and the AIS population. For these two populations, the green AF intensity of both the fingernails and the skin of Dorsal Index Fingers of the smokers was significantly higher, compared with the green AF intensity of the Index Fingernails and the skin of Dorsal Index Fingers of the non-smokers (Figs. 3A and 3B). In contrast, no significant difference was observed between the green AF intensity of the non-smokers and that of the smokers at other examined positions of the skin (Supplemental Figs. 3A, 3B, 3C, 3D, and 3E).

**Fig. 3.**
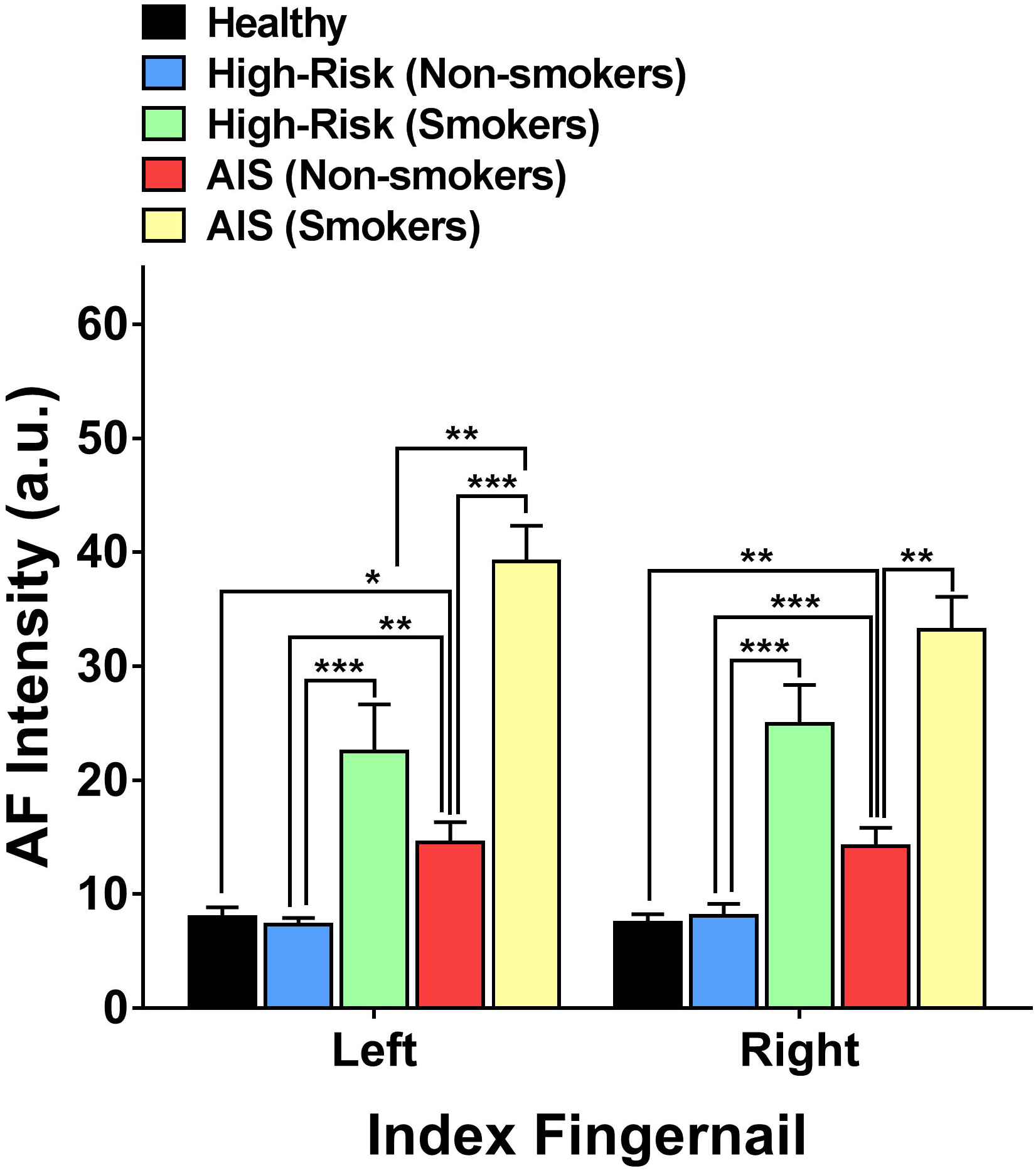
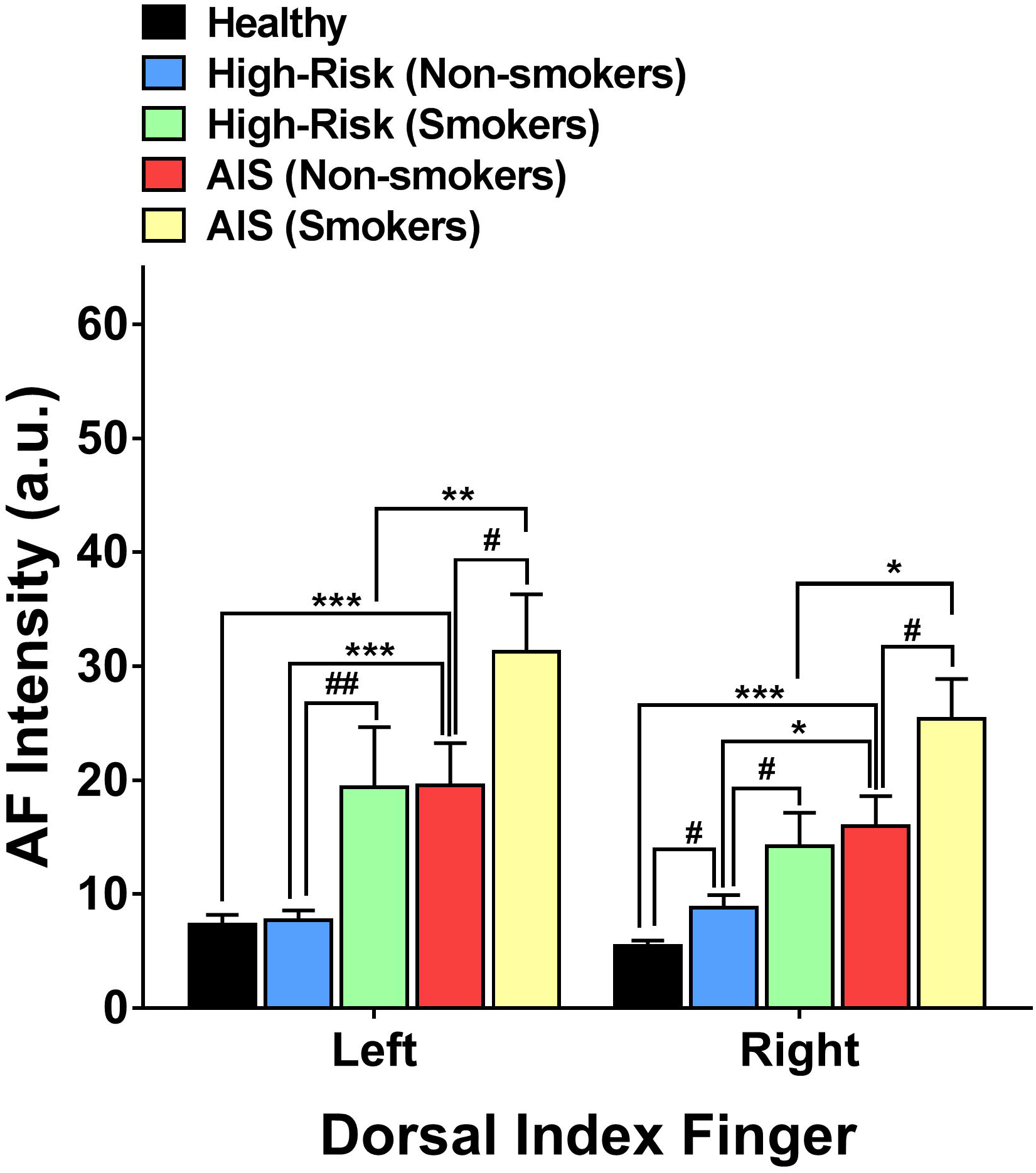
The green AF intensity of the Index Fingernails and the skin of Dorsal Index Fingers of the smokers in both the population at risk of developing stroke and the AIS population was significantly higher than that of the non-smokers. (A) In both right and left Index Fingernails, the AF intensity of the smokers in both the population at risk of developing stroke as well as the AIS population was significantly higher than that of the non-smokers. (B) In the skin of both right and left Dorsal Index Fingers, the AF intensity of the smokers in the population at risk of developing stroke as well as the AIS population was significantly higher than that of the non-smokers. The number of the healthy non-smokers, the non-smokers of the population high-risk of developing stroke, the smokers of the population high-risk of developing stroke, the non-smokers of the AIS population and the smokers of the AIS population was 49-50,76-77, 32-33 39-40 and 39-40, respectively. *, *p* < 0.05; **, *p* < 0.01; ***, *p* < 0.001; #, *p* < 0.05 (Mann-Whitney test); ##, *p* < 0.01 (Mann-Whitney test).

### 3) The green AF asymmetry of the Index Fingernails and the skin of Dorsal Index Fingers of the smokers in both the population high-risk of developing stroke and the AIS population was significantly higher than that of the non-smokers

We also determined the green AF asymmetry of the Index Fingernails and twelve positions of the skin of the population high-risk of developing stroke and the AIS population. For these two populations, the green AF asymmetry of both the fingernails and the skin of Dorsal Index Fingers of the smokers was significantly higher, compared with the green AF intensity of the Index Fingernails and the skin of Dorsal Index Fingers of the non-smokers (Figs. 4A and 4B). In contrast, no significant difference was observed between the green AF asymmetry of the non-smokers and that of the smokers at other examined positions of the skin (Supplemental Figs. 4A, 4B, 4C, 4D, and 4E).

**Fig. 4.**
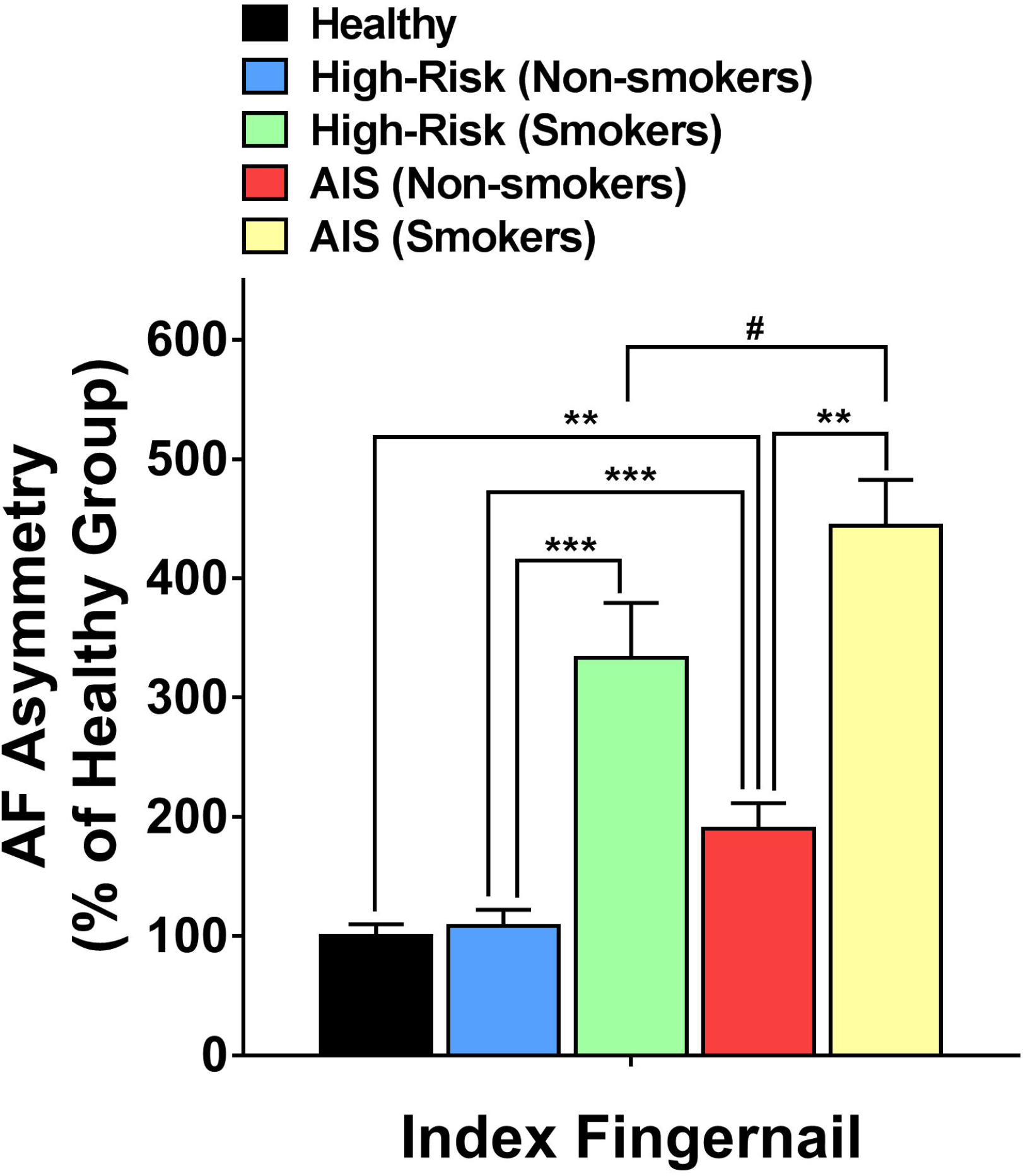
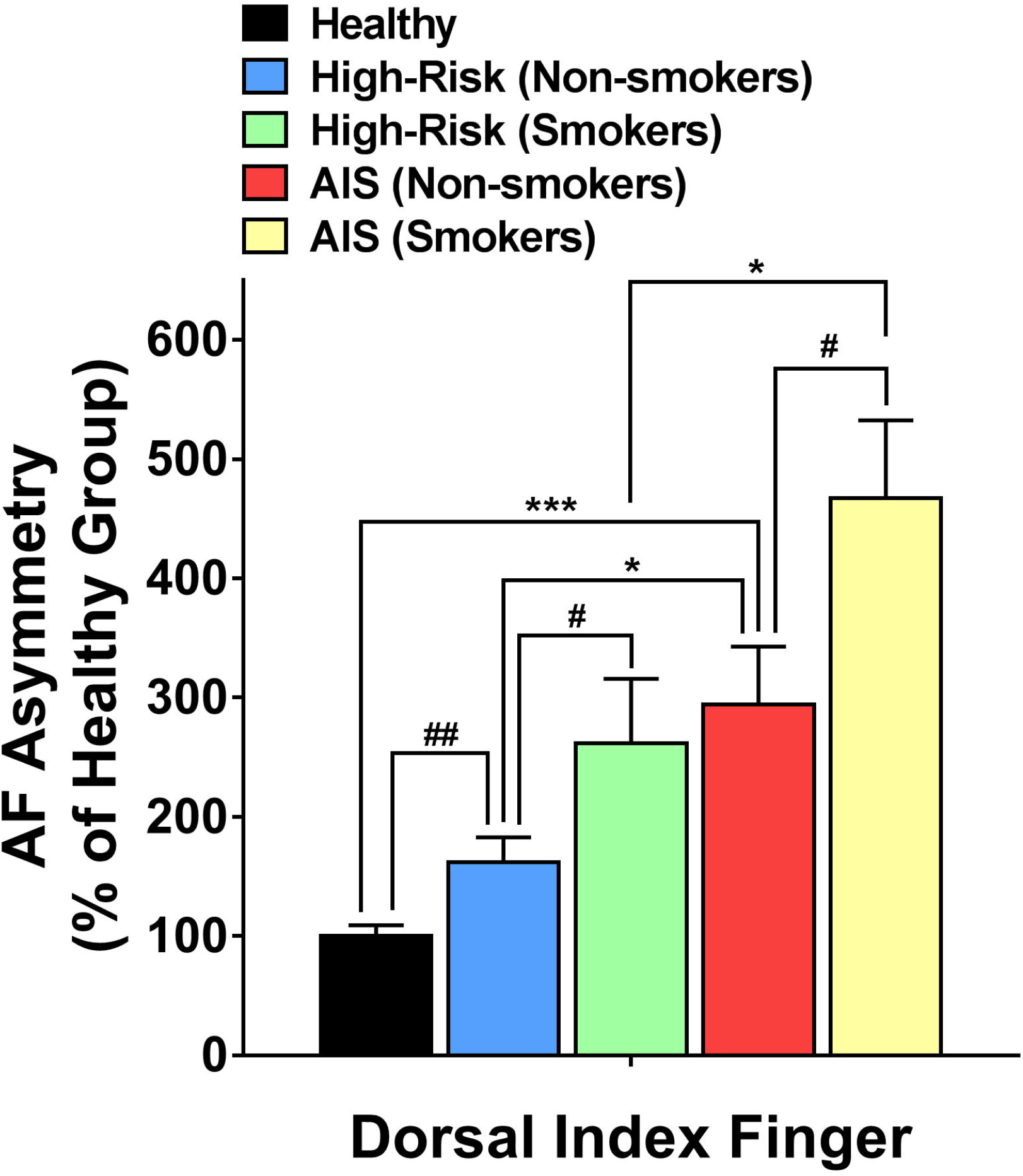
The green AF asymmetry of the Index Fingernails and the skin of Dorsal Index Fingers of the smokers in both the population high-risk of developing stroke and the AIS population was significantly higher than that of the non-smokers. (A) In both right and left Index Fingernails, the AF asymmetry of the smokers in both the population at risk of developing stroke as well as the AIS population was significantly higher than that of the non-smokers. (B) In the skin of both right and left Dorsal Index Fingers, the AF asymmetry of the smokers in the population at risk of developing stroke as well as the AIS population was significantly higher than that of the non-smokers. The number of the healthy non-smokers, the non-smokers of the population high-risk of developing stroke, the smokers of the population high-risk of developing stroke, the non-smokers of the AIS population and the smokers of the AIS population was 49-50,76-77, 32-33 39-40 and 39-40, respectively. *, *p* < 0.05; **, *p* < 0.01; ***, *p* < 0.001; #, *p* < 0.05 (Mann-Whitney test); ##, *p* < 0.01 (Mann-Whitney test).

### 4) The green AF intensity of Index Fingernails and the skin of Dorsal Index Fingers holds great potential for differentiating smokers and non-smokers in natural populations, the population high-risk of developing stroke, and AIS patients

ROC analyses using the green AF intensity of either right or left Index Fingernails showed that the AUC was 0.889 and 0.866, respectively, for differentiating the smokers and the non-smokers in the natural populations at age between 20 - 50 years of old (Fig. 5A). ROC analyses using the green AF intensity of either right or left Dorsal Index Fingers showed that the AUC was 0.819 and 0.774, respectively, for differentiating the smokers and the non-smokers in the natural populations at age between 20 - 50 years of old (Fig. 5B). ROC analyses using the green AF intensity of either right or left Index Fingernails showed that the AUC was 0.825 and 0.864, respectively, for differentiating the smokers and the non-smokers in the natural populations at age between 50 - 80 years of old (Fig. 5C). ROC analyses using the green AF intensity of either right or left Dorsal Index Fingers showed that the AUC was 0.648 and 0.742, respectively, for differentiating the smokers and the non-smokers in the natural populations at age between 50 - 80 years of old (Fig. 5D).

**Fig. 5.**
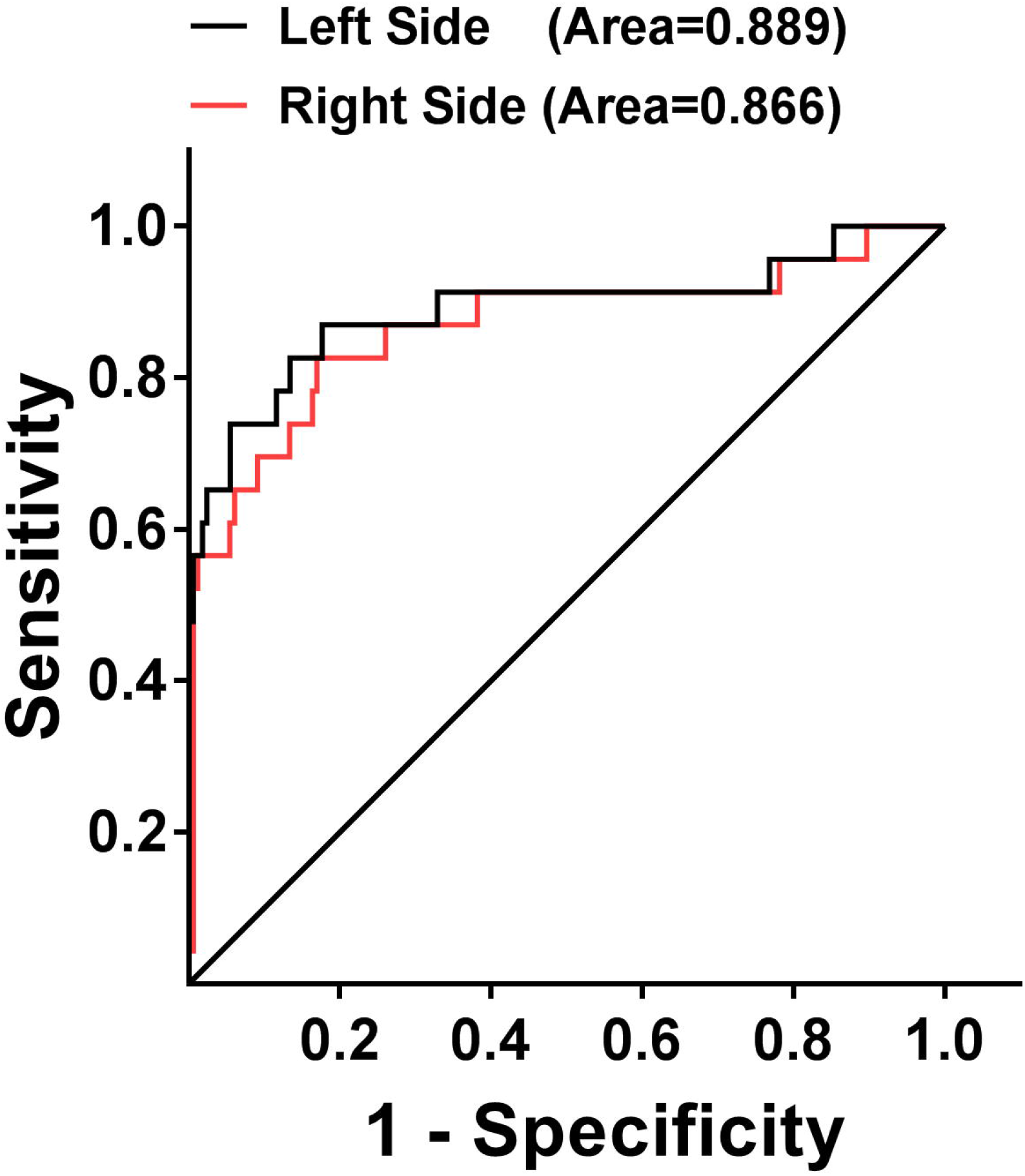
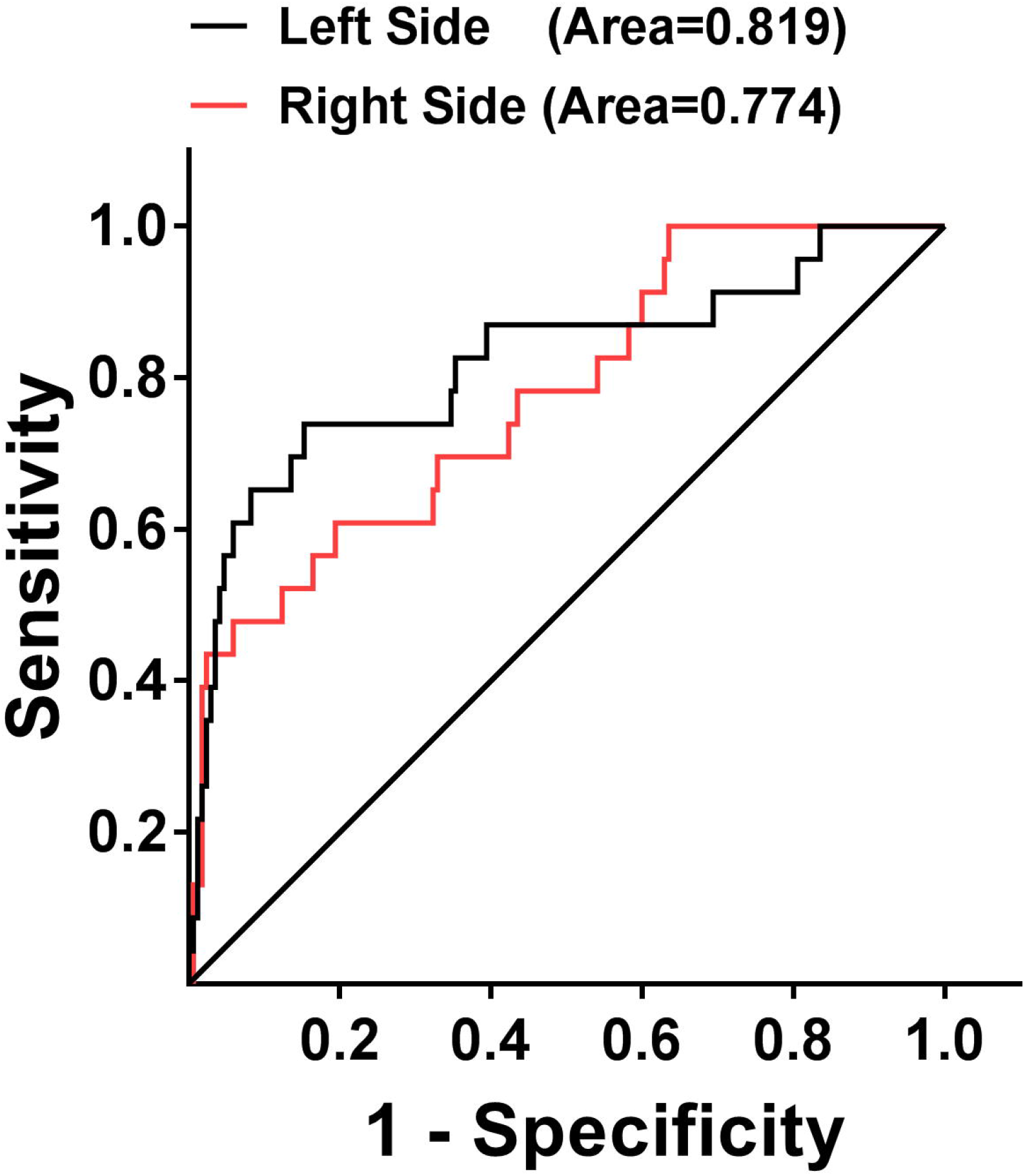
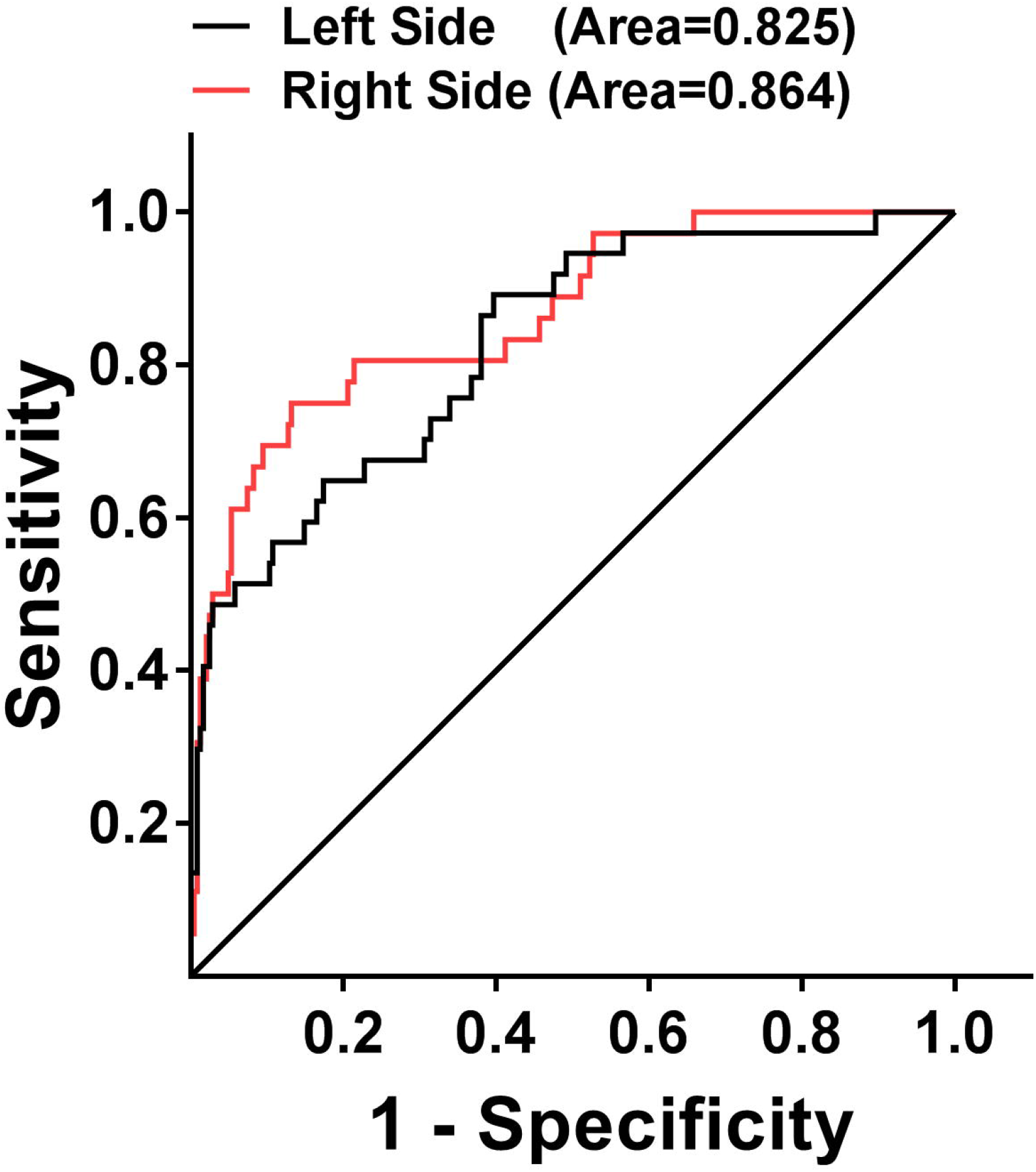
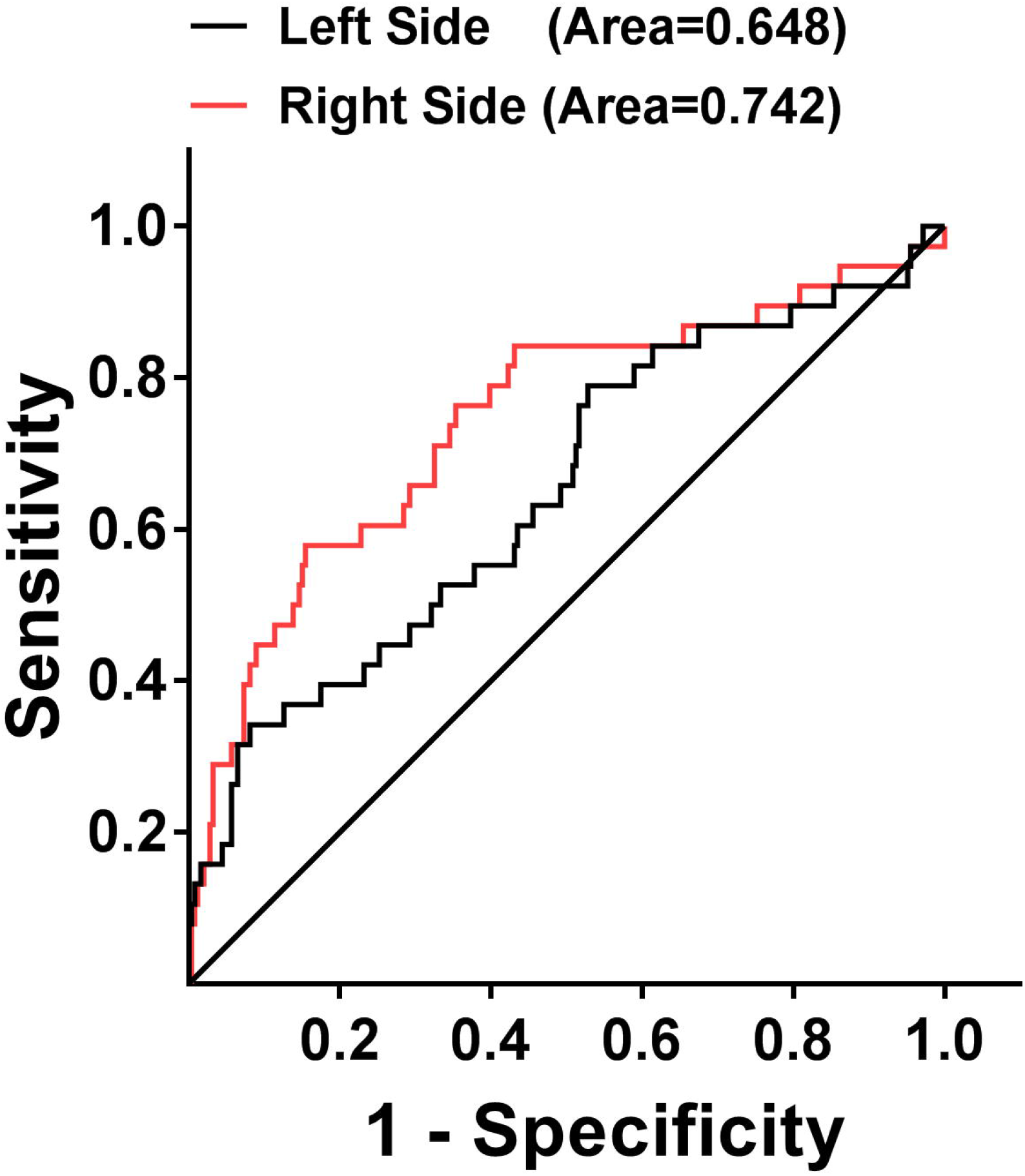
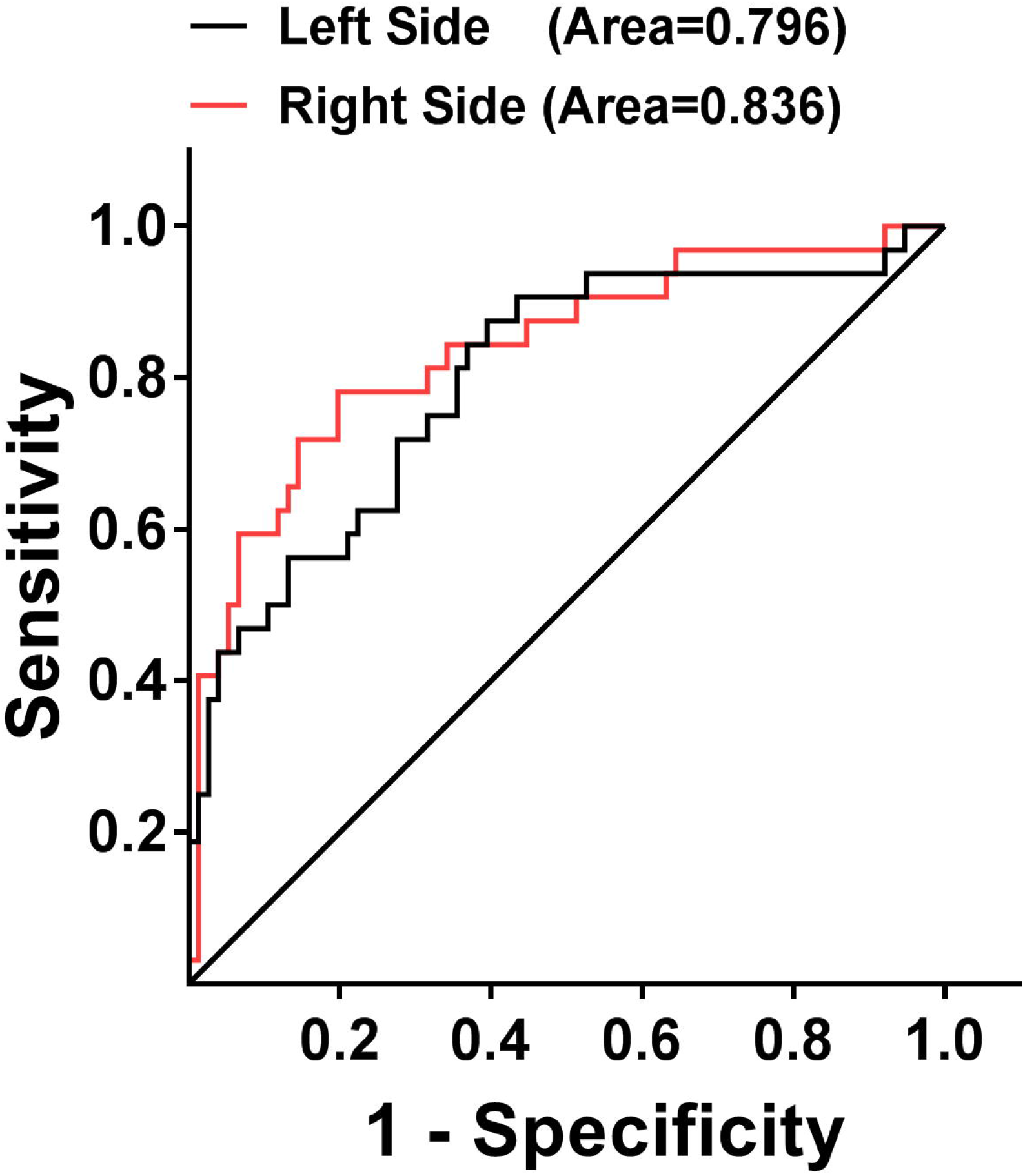
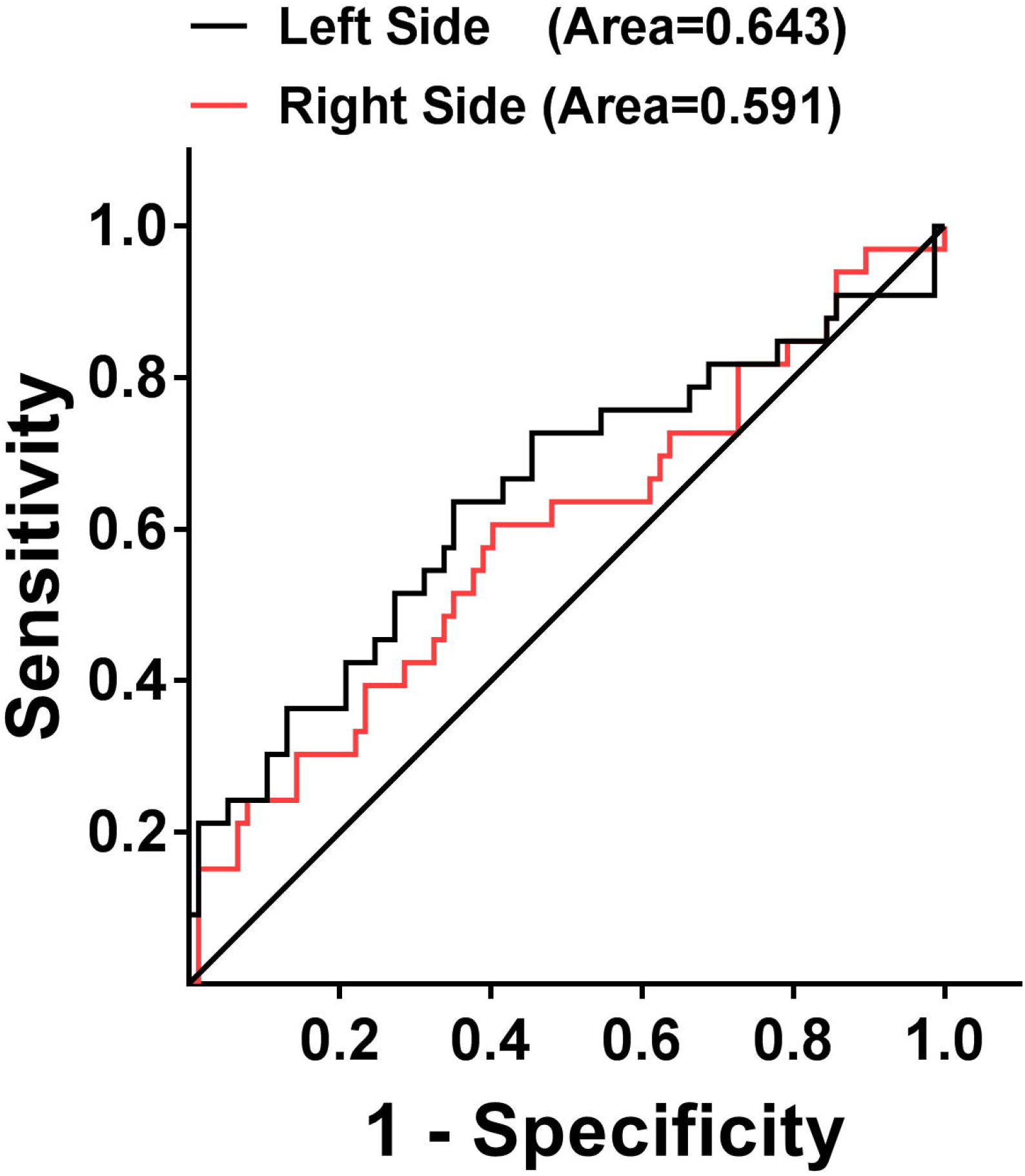
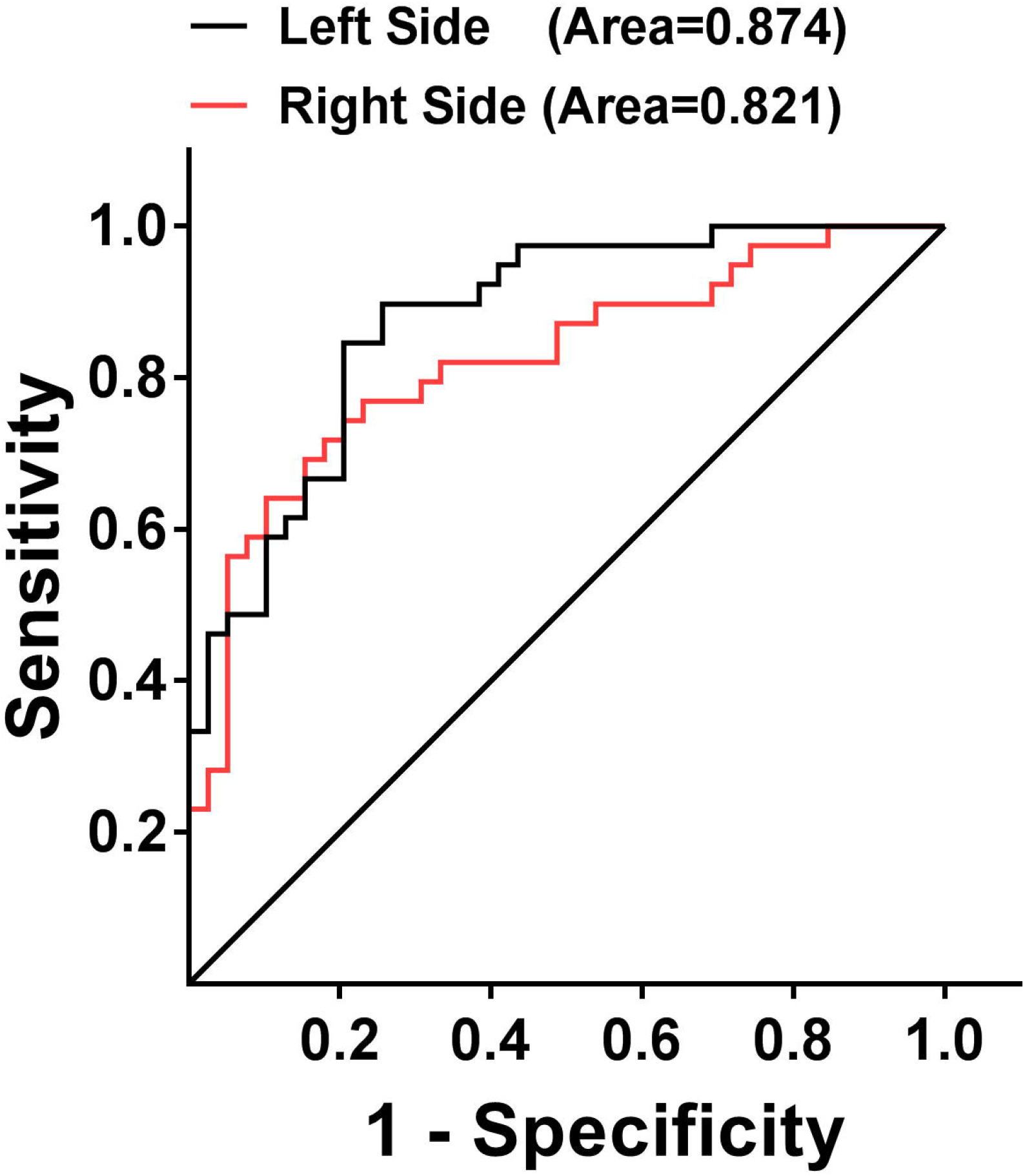
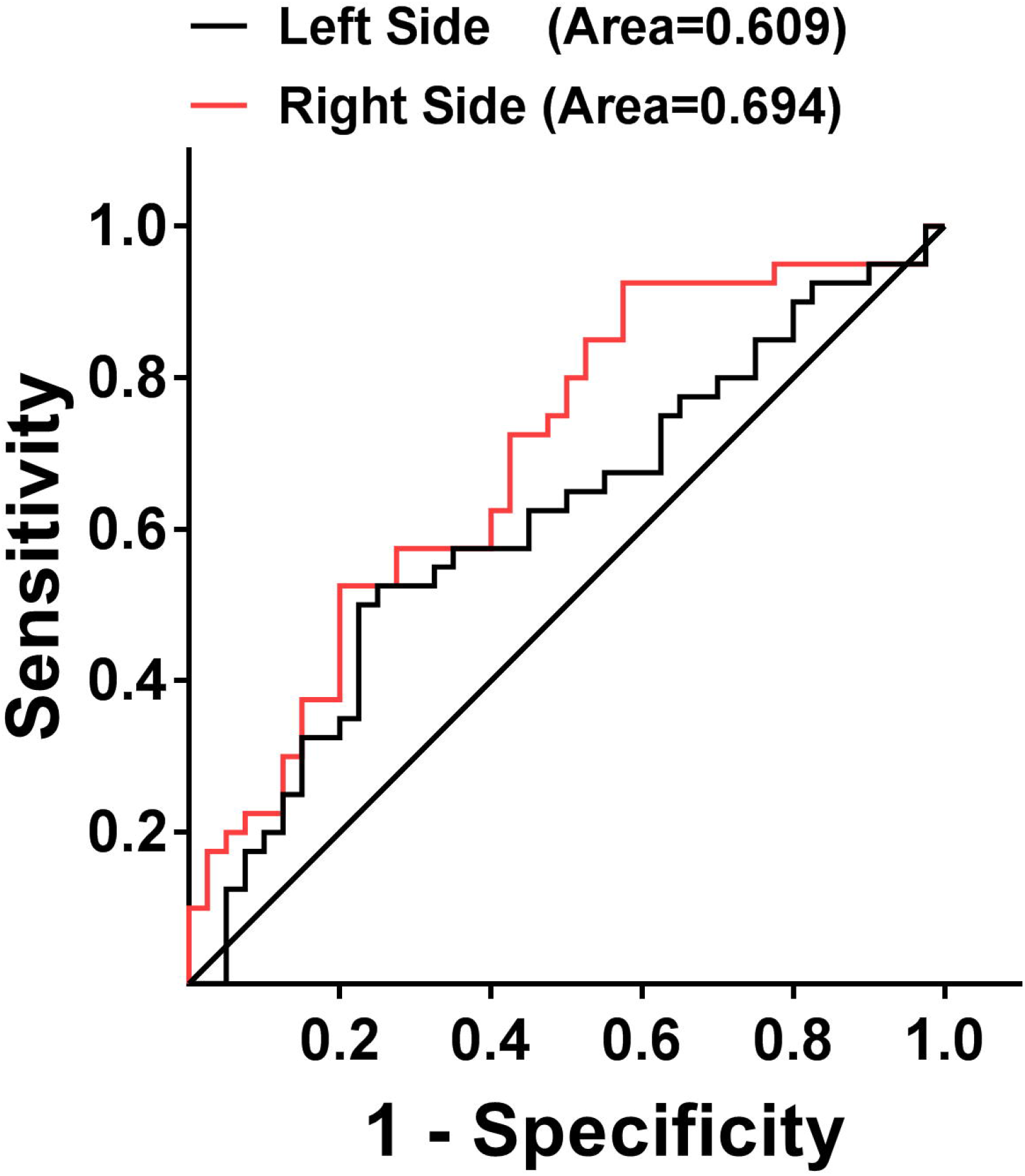
The green AF intensity of Index Fingernails and the skin of Dorsal Index Fingers holds great potential for differentiating the smokers and the non-smokers in natural populations, the population at risk of developing stroke, and AIS patients. (A) ROC analyses using the green AF intensity of either right or left Index Fingernails of the smokers and the non-smokers in the natural population at age between 20 - 50 years of old. (B) ROC analyses using the green AF intensity of either right or left Dorsal Index Fingers of the smokers and the non-smokers in the natural population at age between 20 - 50 years of old. (C) ROC analyses using the green AF intensity of either right or left Index Fingernails of the smokers and the non-smokers in the natural population at age between 50 - 80 years of old. (D) ROC analyses using the green AF intensity of either right or left Dorsal Index Fingers of the smokers and the non-smokers in the natural population at age between 50 - 80 years of old. (E) ROC analyses using the green AF intensity of either right or left Index Fingernails of the smokers and the non-smokers in the population at risk of developing AIS at age between 50 - 80 years of old. (F) ROC analyses using the green AF intensity of either right or left Dorsal Index Fingers of the smokers and the non-smokers in the population at risk of developing AIS at age between 50 - 80 years of old. (G) ROC analyses using the green AF intensity of either right or left Index Fingernails of the smokers and the non-smokers in the AIS population at age between 50 - 80 years of old. (H) ROC analyses using the green AF intensity of either right or left Dorsal Index Fingers of the smokers and the non-smokers in the AIS population at age between 50 - 80 years of old. The number of the non-smokers and the smokers in the age group between 20 - 50 years of old was 165-170 and 23, respectively. The number of the non-smokers and the smokers in the age group between 50 - 80 years of old was 242-246 and 36-38, respectively. The number of the non-smokers of the population high-risk of developing stroke, the smokers of the population high-risk of developing stroke, the non-smokers of the AIS population and the smokers of the AIS population was 76-77, 32-33 39-40 and 39-40, respectively.

ROC analyses using the green AF intensity of either right or left Index Fingernails showed that the AUC was 0.796 and 0.836, respectively, for differentiating the smokers and the non-smokers in the population high-risk of developing stroke (Fig. 5E). ROC analyses using the green AF intensity of the skin of either right or left Dorsal Index Fingers showed that the AUC was 0.643 and 0.591, respectively, for differentiating the smokers and the non-smokers in the population high-risk of developing stroke (Fig. 5F). ROC analyses using the green AF intensity of either right or left Index Fingernails showed that the AUC was 0.874 and 0.821, respectively, for differentiating the smokers and the non-smokers in the AIS population (Fig. 5G). ROC analyses using the green AF intensity of the skin of either right or left Dorsal Index Fingers showed that the AUC was 0.609 and 0.694, respectively, for differentiating the smokers and the non-smokers in the AIS population (Fig. 5H).

## Discussion

The major findings of our current study include: First, for the natural population at age between 20 - 50 years of old, both the green AF intensity and the AF asymmetry of the Index Fingernails and the skin of Dorsal Index Fingers of the smokers were remarkably higher than those of the non-smokers. Second, for the natural population, the population at risk of developing stroke and the AIS population at age between 50 - 80 years of old, both the green AF intensity and the AF asymmetry of the Index Fingernails and the skin of Dorsal Index Fingers of the smokers were also remarkably higher than those of the non-smokers. Third, ROC analyses using the green AF intensity of the Index Fingernails showed that the AUC values were 0.796 to 0.889 for differentiating the smokers and the non-smokers in the natural population, the population at risk of developing stroke and the AIS population. Collectively, our study has indicated that increased green AF intensity of the fingernails and certain locations of the skin is a novel biomarker for smoking.

Tobacco smoking is a major risk factor of multiple major diseases (1-3). It is of critical importance to search for the biomarkers of smoking for non-invasive and rapid monitoring of the pathological changes of smokers’ body. The search is highly valuable for the health monitoring of a large population of smokers, which is a crucial part of health management of the general population. However, there has been no non-invasive method for monitoring the health state of smokers.

Our current study has provided evidence indicating that the green AF intensity of the Index Fingernails and the skin of certain locations such as Dorsal Index Fingers is a novel biomarker for smokers of natural populations, the population at risk of developing stroke, and AIS patients: ROC analyses using the green AF intensity of these positions showed highly promising AUC values for differentiating the smokers and the non-smokers in these populations. Based on this finding, approaches for non-invasive, economic and efficient monitoring the heathy state of the smokers may be established, which may generate tremendous health benefits. With future studies on the AF of the skin and fingernails of smokers and non-smokers as well as applications of AI technology, it is expected that increasingly precise evaluations on the smokers’ health status could be established.

Our previous studies have suggested that the ‘Pattern of AF’ is a novel biomarker that can be used for non-invasive diagnosis of several major diseases, which forms a basis for establishing a new biomedical imaging technology (10-12). In order to improve this new technology, it is critical to further investigate the factors that can influence the AF in natural populations, populations high-risk of developing major diseases as well as patient populations. Our current study has indicated that smoking is an important factor that is associated with increased AF intensity and AF asymmetry of the fingernails and certain positions of the skin in these populations. These findings are highly valuable for understanding the mechanisms underlying the changes of the AF in these populations, which is required for establishing the AF-based models for disease diagnosis and evaluations of the health conditions of natural populations.

Our previous study has shown that both inflammation and oxidative stress can dose-dependently increase the epidermal green AF (10). It is established that smoking can cause increased inflammation and oxidative stress in human body (8, 9, 13, 14). Therefore, we propose that the capacity of smoking to increase the AF in these populations may result from the capacity of smoking to increase inflammation and oxidative stress in the body of these populations.

It is noteworthy that in these populations, smoking was selectively associated with increased AF intensity of the fingernails and the skin of Dorsal Index Fingers. Since it takes a significant duration of time for the growth of nails, we propose that increased AF of the nails is a biomarker for the levels of chronic exposures of the nails to inflammation and oxidative stress in the body. Based on this proposal, it is reasonable to observe the increased AF in the fingernails of the people in these populations. Future studies are required to investigate the mechanisms underlying the AF of the nails.

Based on the observations of our previous studies, we have proposed that the positions with increased AF in the skin are associated with the ‘channels’ (or called Meridian) in traditional Chinese medicine. According to the Meridian theory, the skin of Dorsal Index Fingers is close to the Large Intestine Meridian. Therefore, our finding that smoking increases the AF of the skin of Dorsal Index Fingers has implicated that smoking may increase the inflammation and oxidative stress of the organs that are associated with the Large Intestine Meridian.

Our study has found that smoking can enhance not only the green AF intensity but also the AF asymmetry of the fingernails and certain positions of the skin in these populations. Our previous study has indicated that high levels of AF asymmetry are associated with major vascular diseases such as AIS (11). Therefore, we propose that smoking may have particularly strong capacity to increase vascular damage, which is consistent with previous reports (9).

## Data Availability

All data referred to in the manuscript are available.

## Acknowledgment

The authors would like to acknowledge the financial support by two research grants from a Major Special Program Grant of Shanghai Municipality (Grant # 2017SHZDZX01) (to W.Y.).

## Legends of Supplemental Figures

**Supplemental Fig. 1. The green AF intensity of the skin at other examined positions of the smokers in natural populations was not significantly different from that of the non-smokers**. The number of the non-smokers and the smokers in the age group between 20 - 50 years of old was 165-170 and 23, respectively. The number of the non-smokers and the smokers in the age group between 50 - 80 years of old was 242-246 and 36-38, respectively. ***, *p* < 0.001.

**Supplemental Fig. 2. The AF asymmetry of the skin at other examined positions of the smokers in natural populations was not significantly different from that of the non-smokers**. The number of the non-smokers and the smokers in the age group between 20 - 50 years of old was 165-170 and 23, respectively. The number of the non-smokers and the smokers in the age group between 50 - 80 years of old was 243-246 and 36-38, respectively. ***, *p* < 0.001.

**Supplemental Fig. 3. The green AF intensity of the skin of at other examined positions of the smokers in the population at risk of developing AIS and the AIS population was not significantly different from that of the non-smokers**. The number of the healthy non-smokers, the non-smokers of the population high-risk of developing stroke, the smokers of the population high-risk of developing stroke, the non-smokers of the AIS population and the smokers of the AIS population was 49-50,76-77, 32-33 39-40 and 39-40, respectively. *, *p* < 0.05; **, *p* < 0.01; ***, *p* < 0.001; #, *p* < 0.05 (Mann-Whitney test); ##, *p* < 0.01 (Mann-Whitney test); ###, *p* < 0.001 (Mann-Whitney test).

**Supplemental Fig. 4. The AF asymmetry of the skin at other examined positions of the smokers in the population at risk of developing AIS and the AIS population was not significantly different from that of the non-smokers**. The number of the healthy non-smokers, the non-smokers of the population high-risk of developing stroke, the smokers of the population high-risk of developing stroke, the non-smokers of the AIS population and the smokers of the AIS population was 49-50,76-77, 32-33 39-40 and 39-40, respectively. *, *p* < 0.05; ***, *p* < 0.001; #, *p* < 0.05 (Mann-Whitney test); ##, *p* < 0.01 (Mann-Whitney test); ###, *p* < 0.001 (Mann-Whitney test).

## Notes

### Competing Interest Statement

The authors have declared no competing interest.

### Funding Statement

Supported by two research grants from a Major Special Program Grant of Shanghai Municipality (Grant # 2017SHZDZX01) and a Major Research Grant from the Scientific Committee of Shanghai Municipality #16JC1400502.

### Author Declarations

The study was conducted according to a protocol approved by the Ethics Committee of Shanghai Fifth People Hospital affiliated to Fudan University.

### Summary of Updates

The following new information been included in this revised article: 1) The comparisons of the green autofluorescence of the Index Fingernails and Dorsal Index Fingers of the non-smokers and smokers of the natural population at age between 20 - 50. 2) We have conducted comparisons of the green autofluorescence of the Index Fingernails and Dorsal Index Fingers of the non-smokers and that of the smokers of the populations at risk of developing stroke and the stroke population.

## References

1. Larsson, S. C., Burgess, S., and Michaelsson, K. (2019) Smoking and stroke: A mendelian randomization study. Ann Neurol 86, 468–471

2. Markidan, J., Cole, J. W., Cronin, C. A., Merino, J. G., Phipps, M. S., Wozniak, M. A., and Kittner, S. J. (2018) Smoking and Risk of Ischemic Stroke in Young Men. Stroke 49, 1276–1278

3. Ambrose, J. A., and Barua, R. S. (2004) The pathophysiology of cigarette smoking and cardiovascular disease: an update. J Am Coll Cardiol 43, 1731–1737

4. Sasco, A. J., Secretan, M. B., and Straif, K. (2004) Tobacco smoking and cancer: a brief review of recent epidemiological evidence. Lung Cancer 45 Suppl 2, S3–9

5. Moran, C., Munch, G., Forbes, J. M., Beare, R., Blizzard, L., Venn, A. J., Phan, T. G., Chen, J., and Srikanth, V. (2015) Type 2 diabetes, skin autofluorescence, and brain atrophy. Diabetes 64, 279–283

6. Pena, A., Strupler, M., Boulesteix, T., and Schanne-Klein, M. (2005) Spectroscopic analysis of keratin endogenous signal for skin multiphoton microscopy. Opt Express 13, 6268–6274

7. Bader, A. N., Pena, A. M., Johan van Voskuilen, C., Palero, J. A., Leroy, F., Colonna, A., and Gerritsen, H. C. (2011) Fast nonlinear spectral microscopy of in vivo human skin. Biomed Opt Express 2, 365–373

8. Dikalov, S., Itani, H., Richmond, B., Vergeade, A., Rahman, S. M. J., Boutaud, O., Blackwell, T., Massion, P. P., Harrison, D. G., and Dikalova, A. (2019) Tobacco smoking induces cardiovascular mitochondrial oxidative stress, promotes endothelial dysfunction, and enhances hypertension. Am J Physiol Heart Circ Physiol 316, H639–H646

9. Siasos, G., Tsigkou, V., Kokkou, E., Oikonomou, E., Vavuranakis, M., Vlachopoulos, C., Verveniotis, A., Limperi, M., Genimata, V., Papavassiliou, G., Stefanadis, C., and Tousoulis, D. (2014) Smoking and atherosclerosis: mechanisms of disease and new therapeutic approaches. Curr Med Chem 21, 3936–3948

10. Zhang, M, Li, Y, Maharjan D. T., He, H., Tao, Y., Wu, D., Ying, W. (2019) Keratin-based epidermal green autofluorescence is a common biomarker of organ injury. bioRxiv 564112

11. Wu, D, Zhang, M., Tao, Y., Li, Y., Zhang, S., Chen, X., Ying, W. (2018) Distinct pattern of autofluorescence of the skin and fingernails of acute ischemic stroke patients: A novel diagnostic biomarker for acute ischemic stroke. bioRxiv, 310904

12. Zhang, M., Tao, Y., Chang, Q., Li, Y., Chu, T., Ying, W. (2020) Selectively increased autofluorescence at certain locations of skin may become a novel diagnostic biomarker for lung cancer. bioRxiv 315440

13. McEvoy, J. W., Nasir, K., DeFilippis, A. P., Lima, J. A., Bluemke, D. A., Hundley, W. G., Barr, R. G., Budoff, M. J., Szklo, M., Navas-Acien, A., Polak, J. F., Blumenthal, R. S., Post, W. S., and Blaha, M. J. (2015) Relationship of cigarette smoking with inflammation and subclinical vascular disease: the Multi-Ethnic Study of Atherosclerosis. Arterioscler Thromb Vasc Biol 35, 1002–1010

14. Rom, O., Avezov, K., Aizenbud, D., and Reznick, A. Z. (2013) Cigarette smoking and inflammation revisited. Respir Physiol Neurobiol 187, 5–10

