## Supplementary figures and images for "Smoking is associated with significantly increased green autofluorescence intensity and asymmetry of the skin and the fingernails of natural populations, population high-risk of developing stroke, and population of acute ischemic stroke"

### Supplemental Fig. 1A

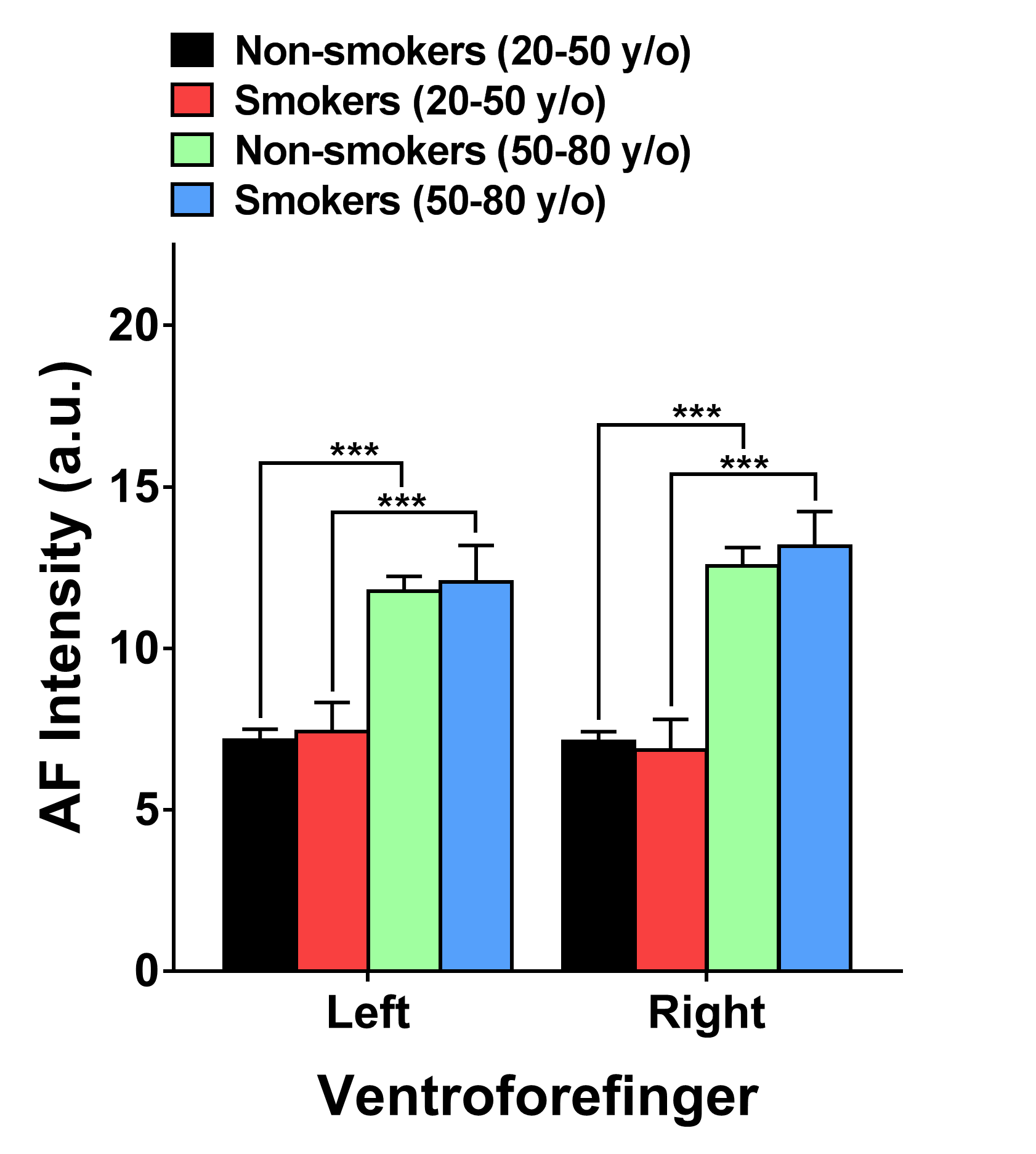

### Supplemental Fig. 1B

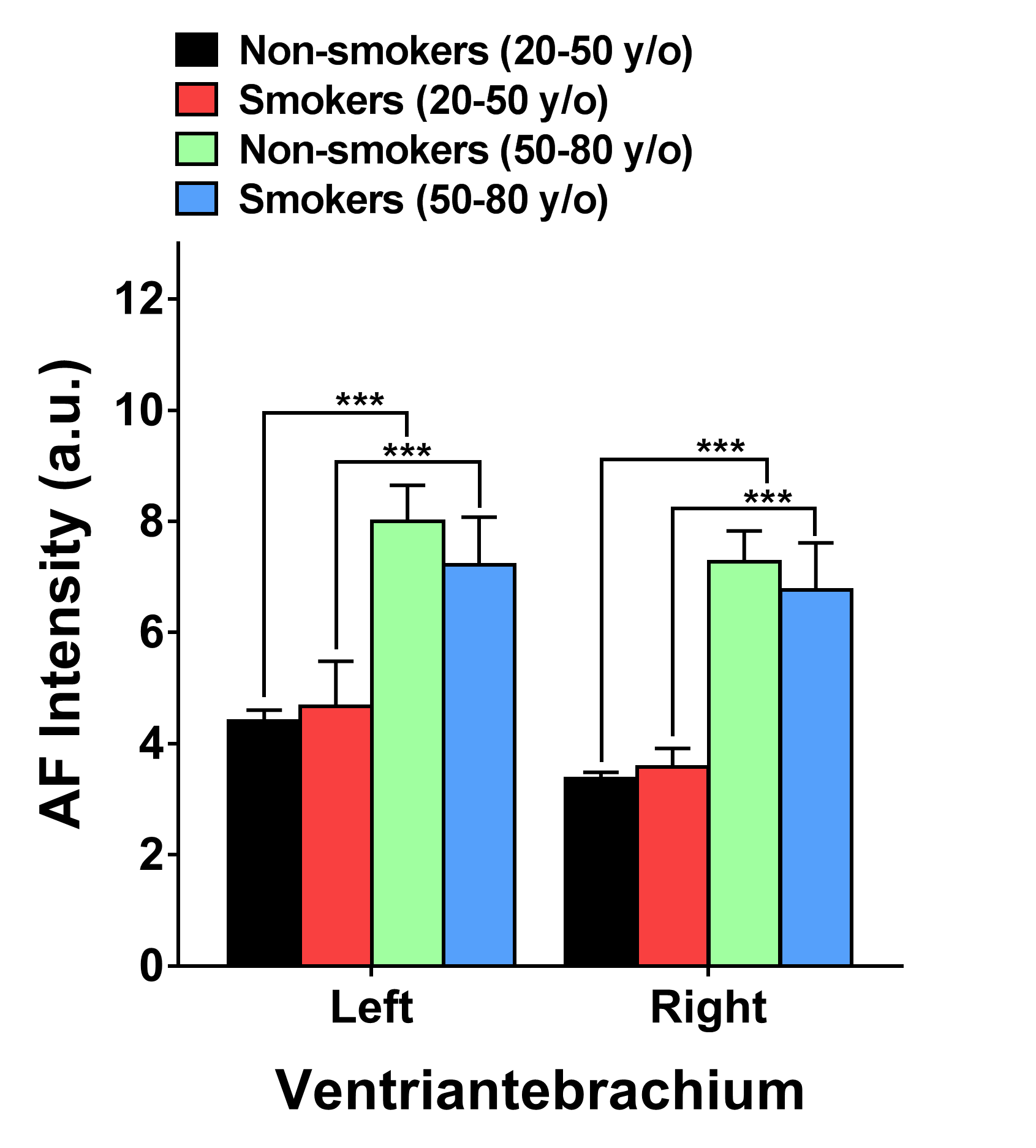

### Supplemental Fig. 1C

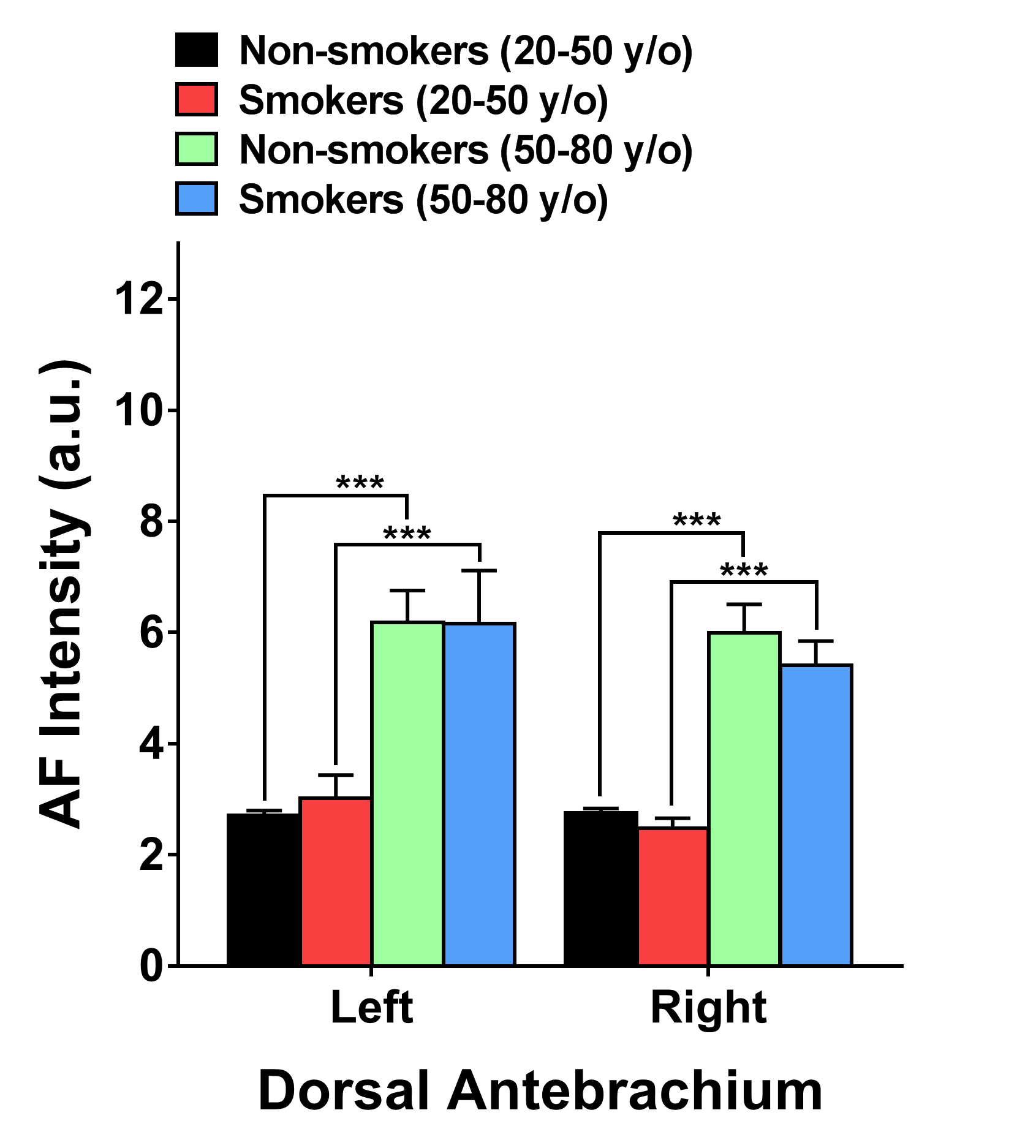

### Supplemental Fig. 1D

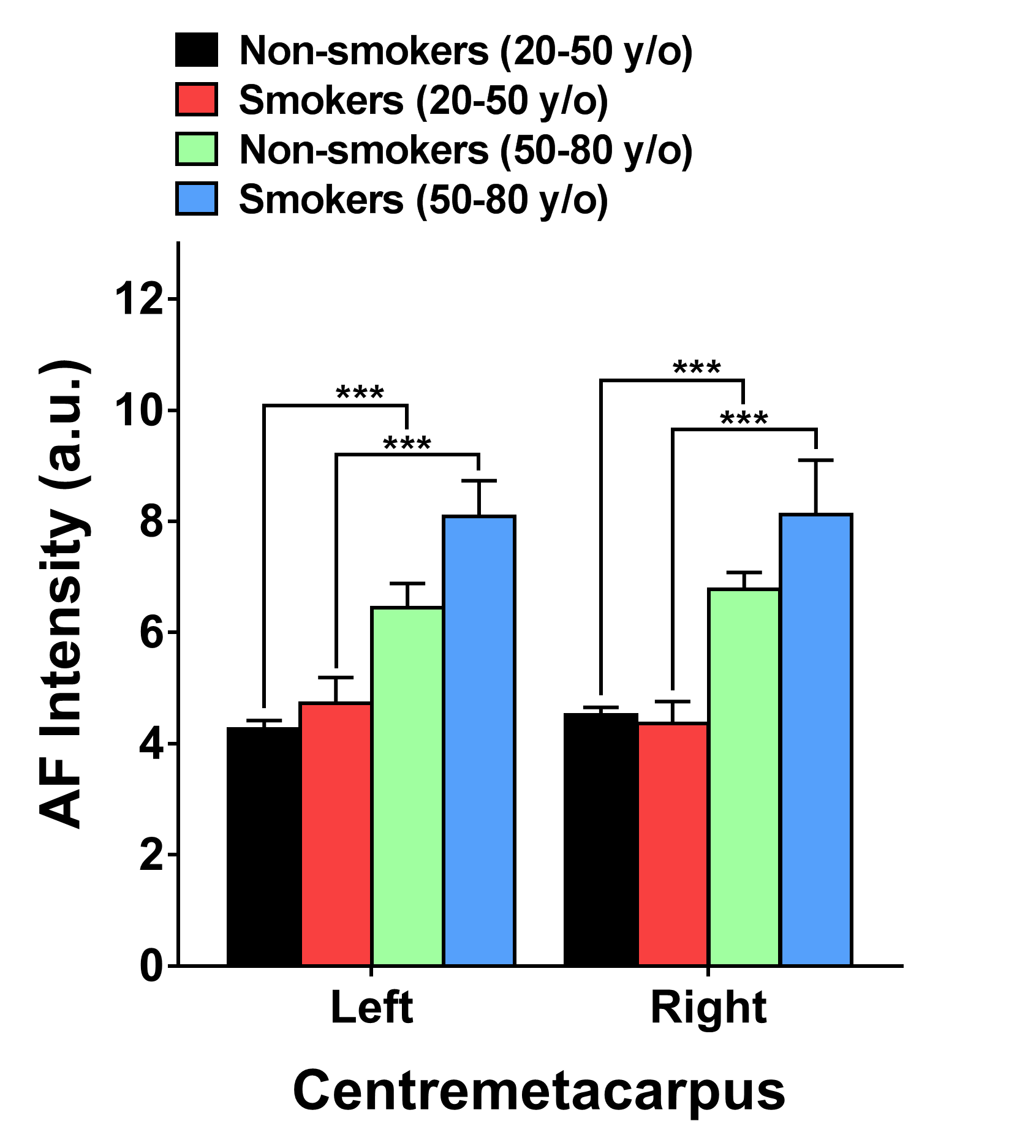

### Supplemental Fig. 1E

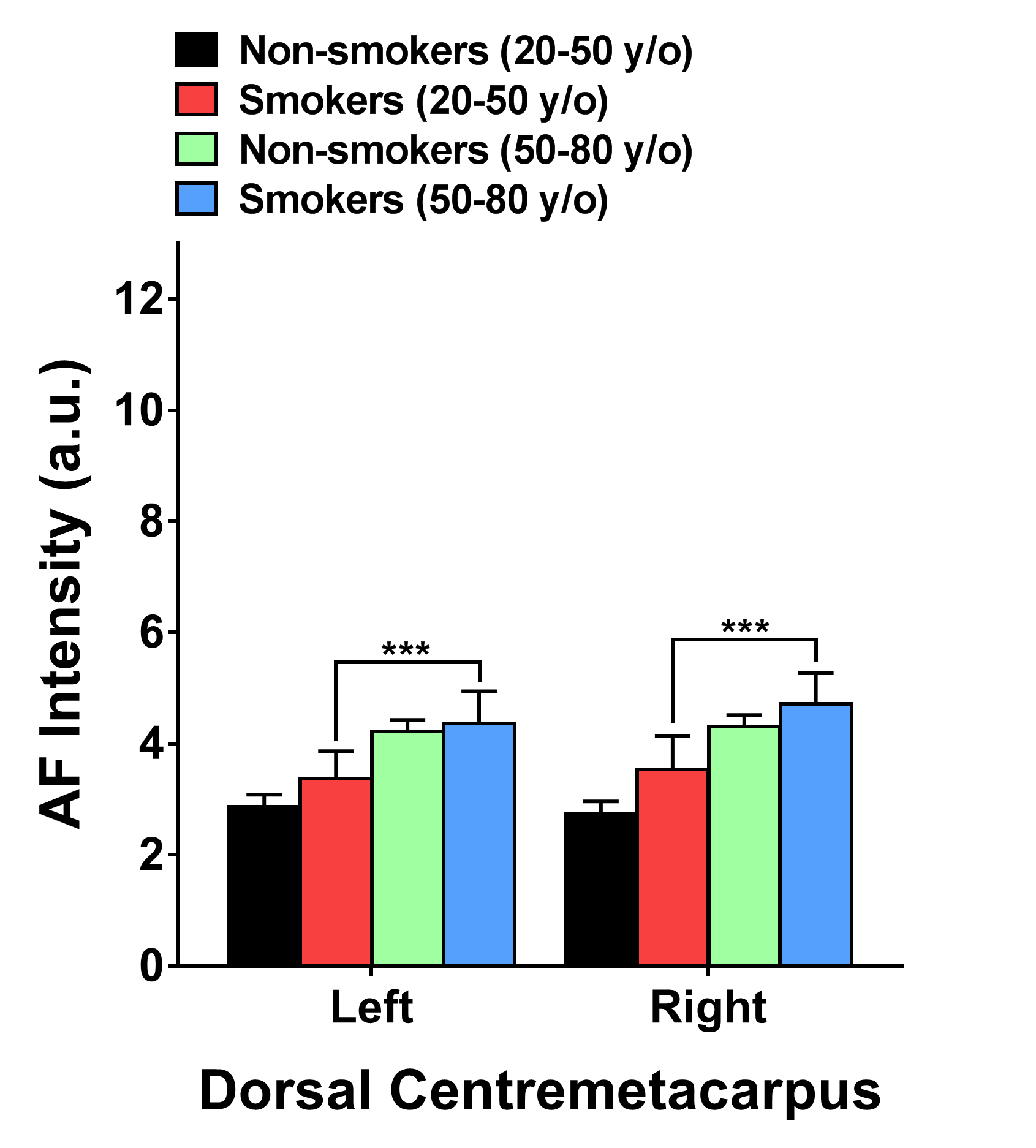

### Supplemental Fig. 2A

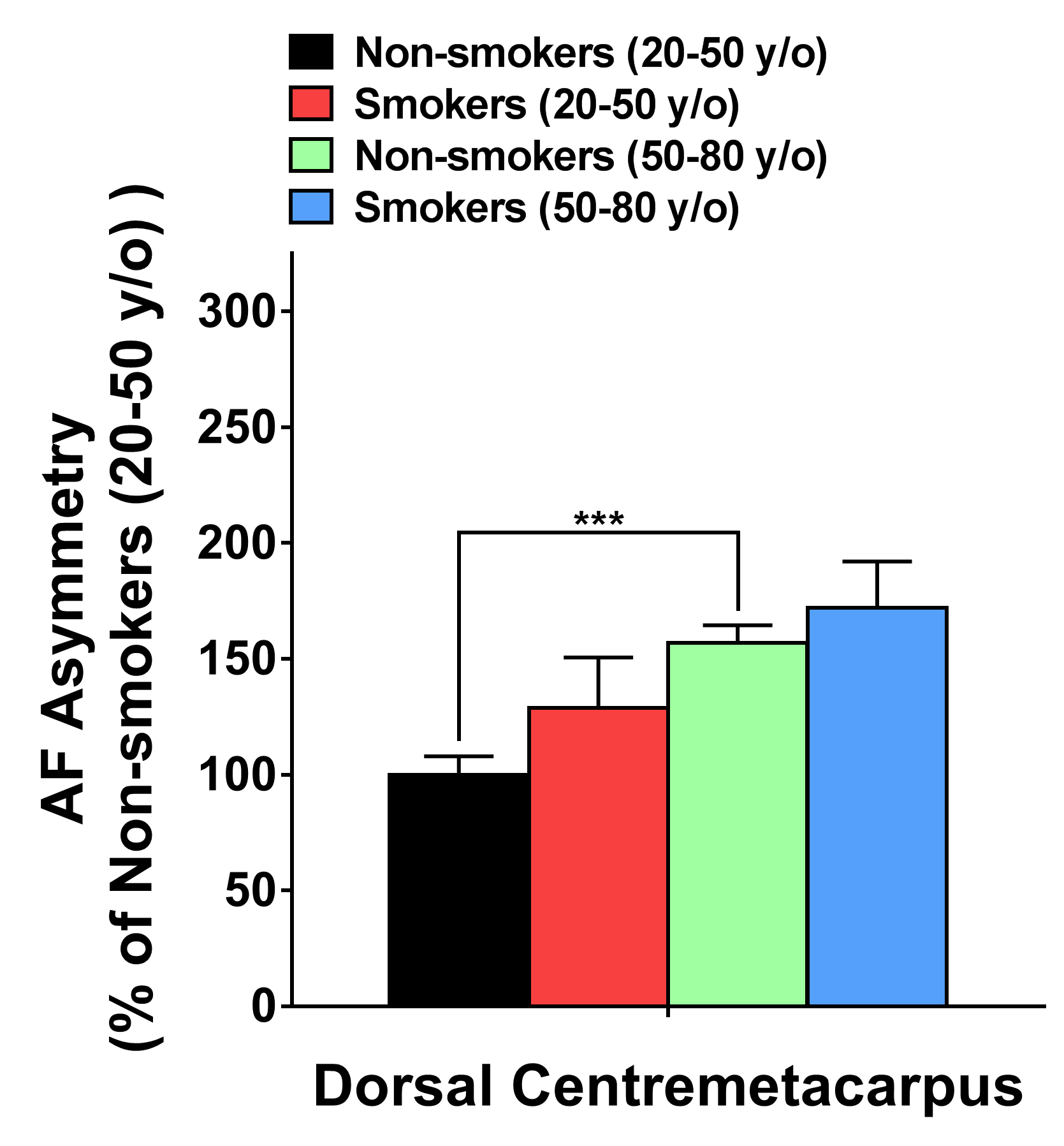

### Supplemental Fig. 2B

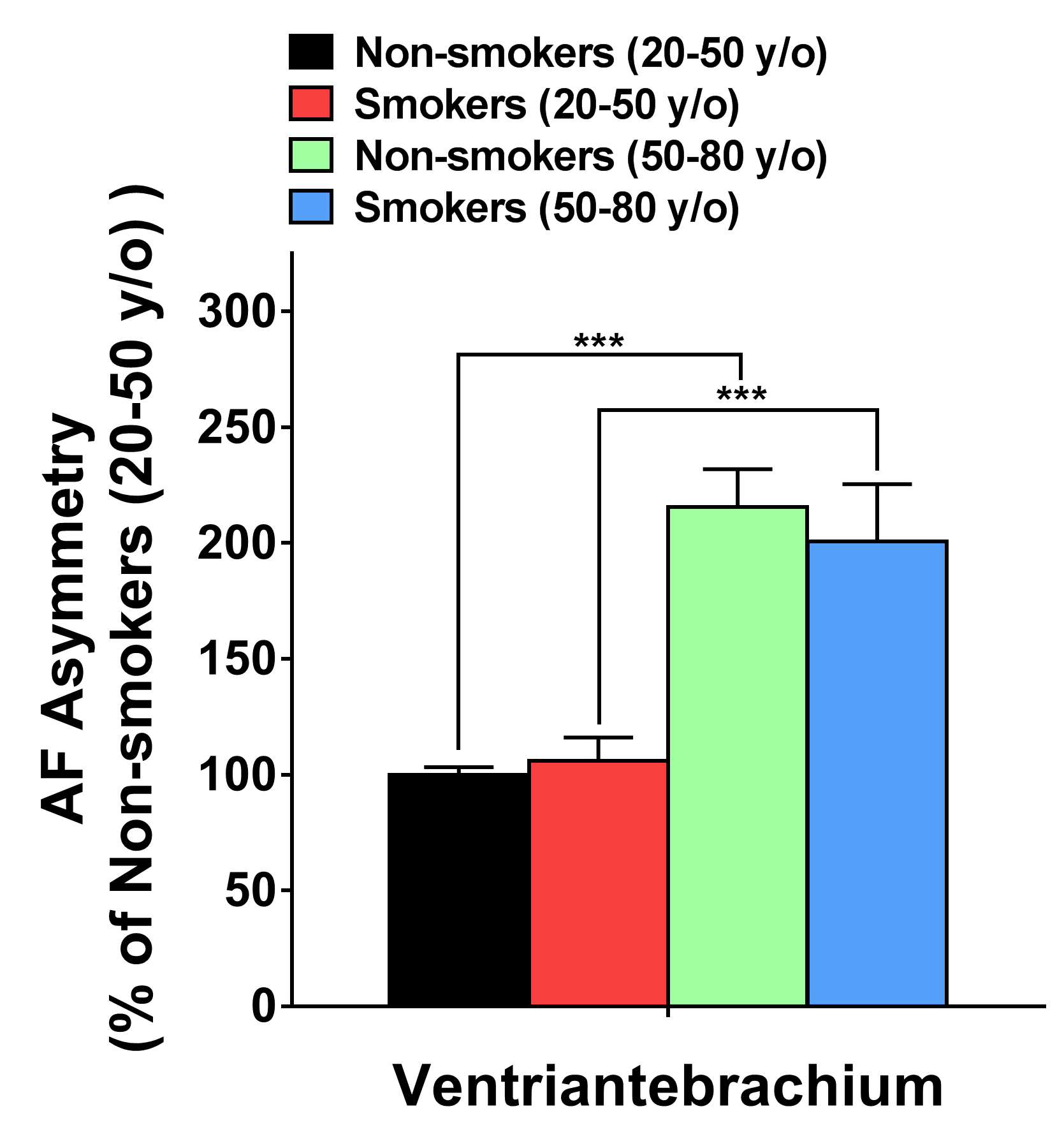

### Supplemental Fig. 2C

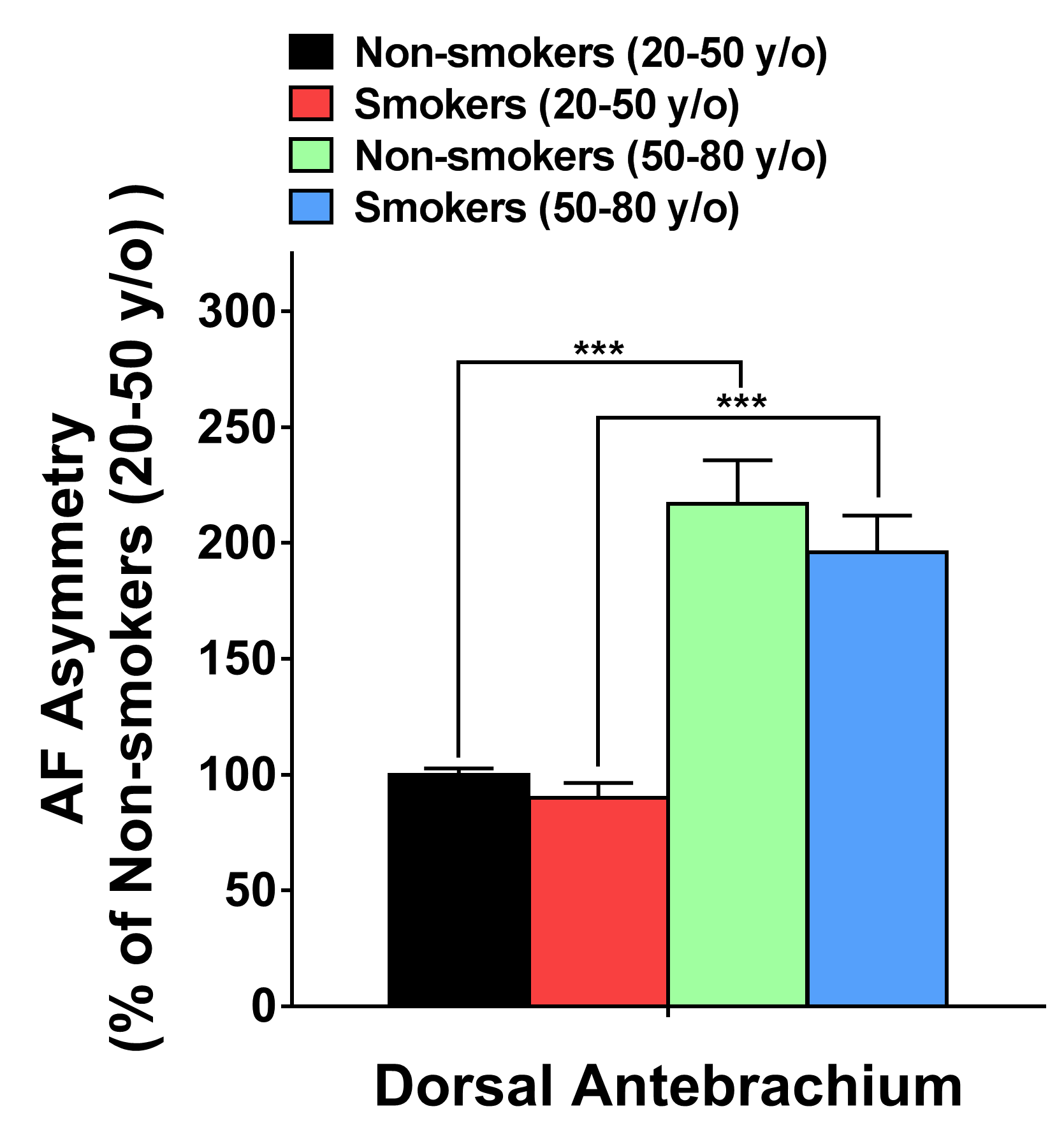

### Supplemental Fig. 2D

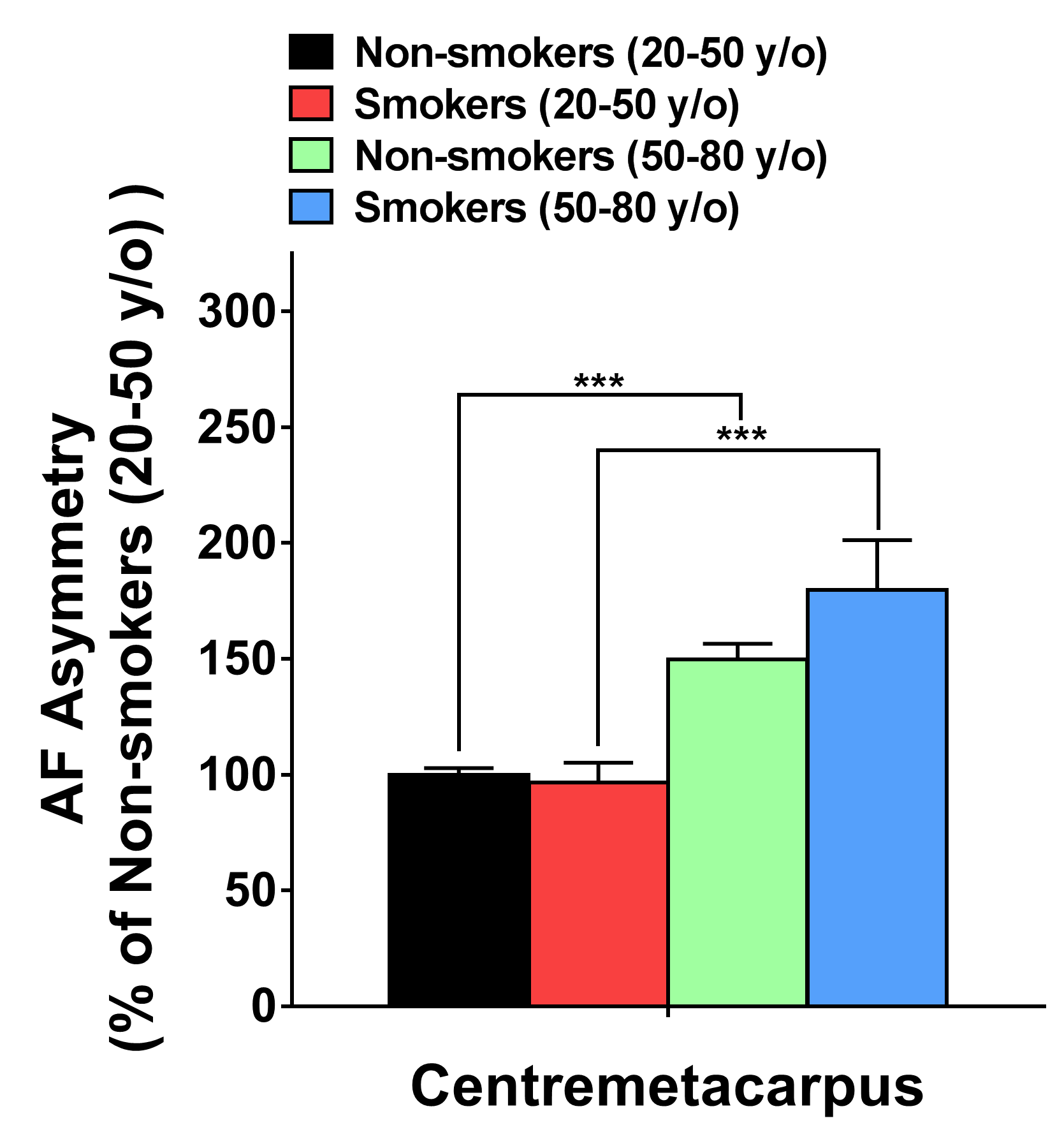

### Supplemental Fig. 3A

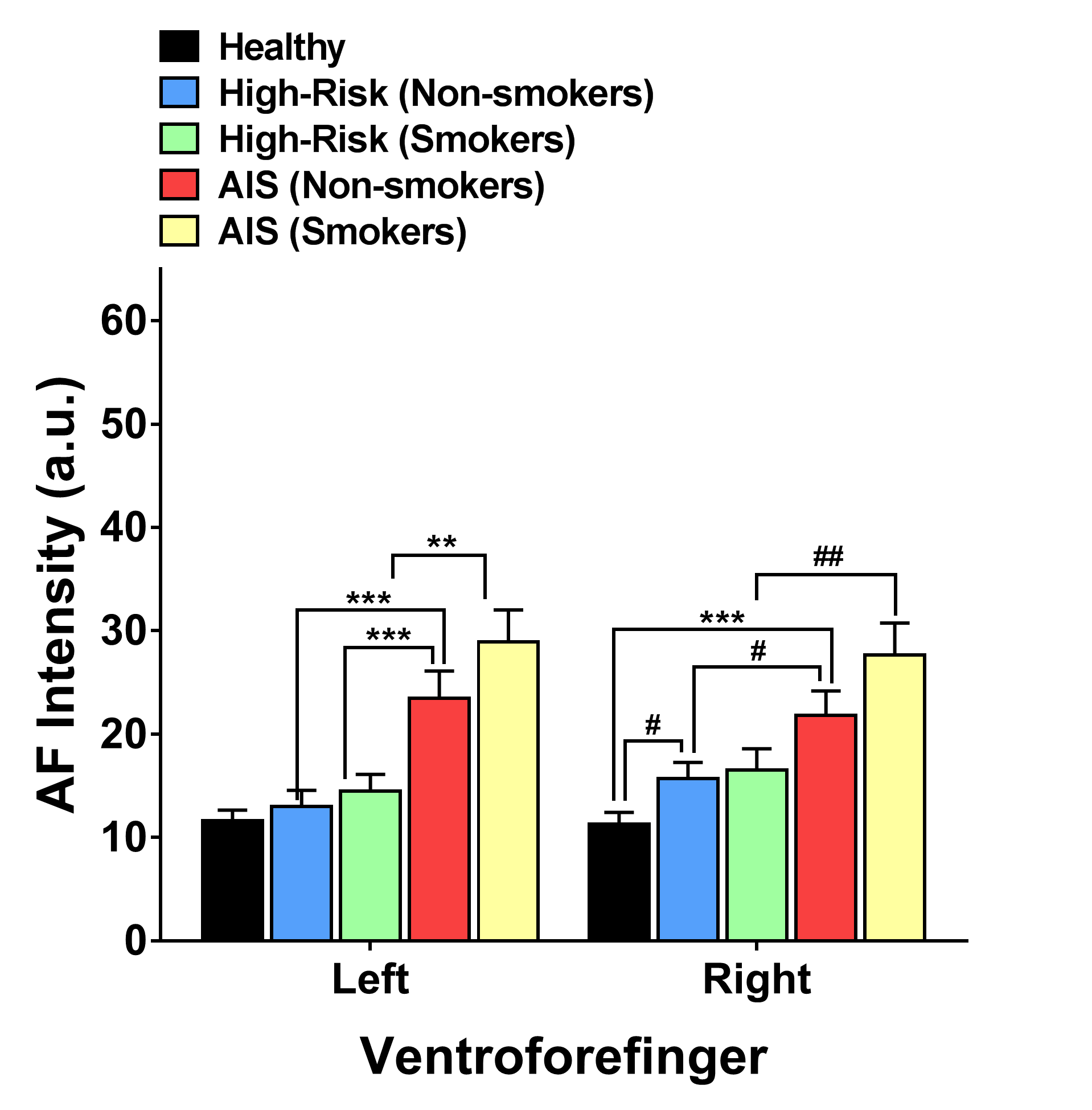

### Supplemental Fig. 3B

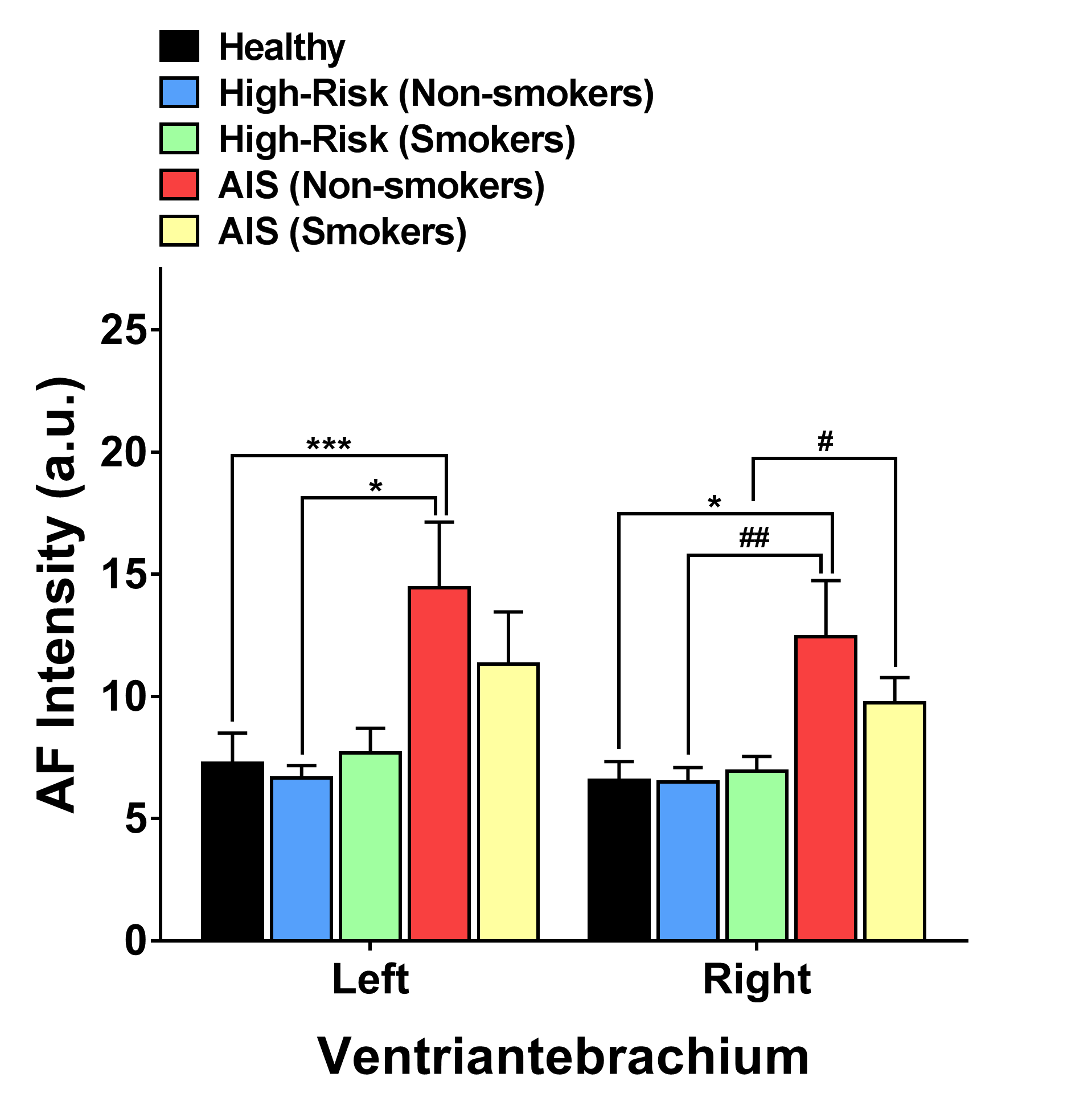

### Supplemental Fig. 3C

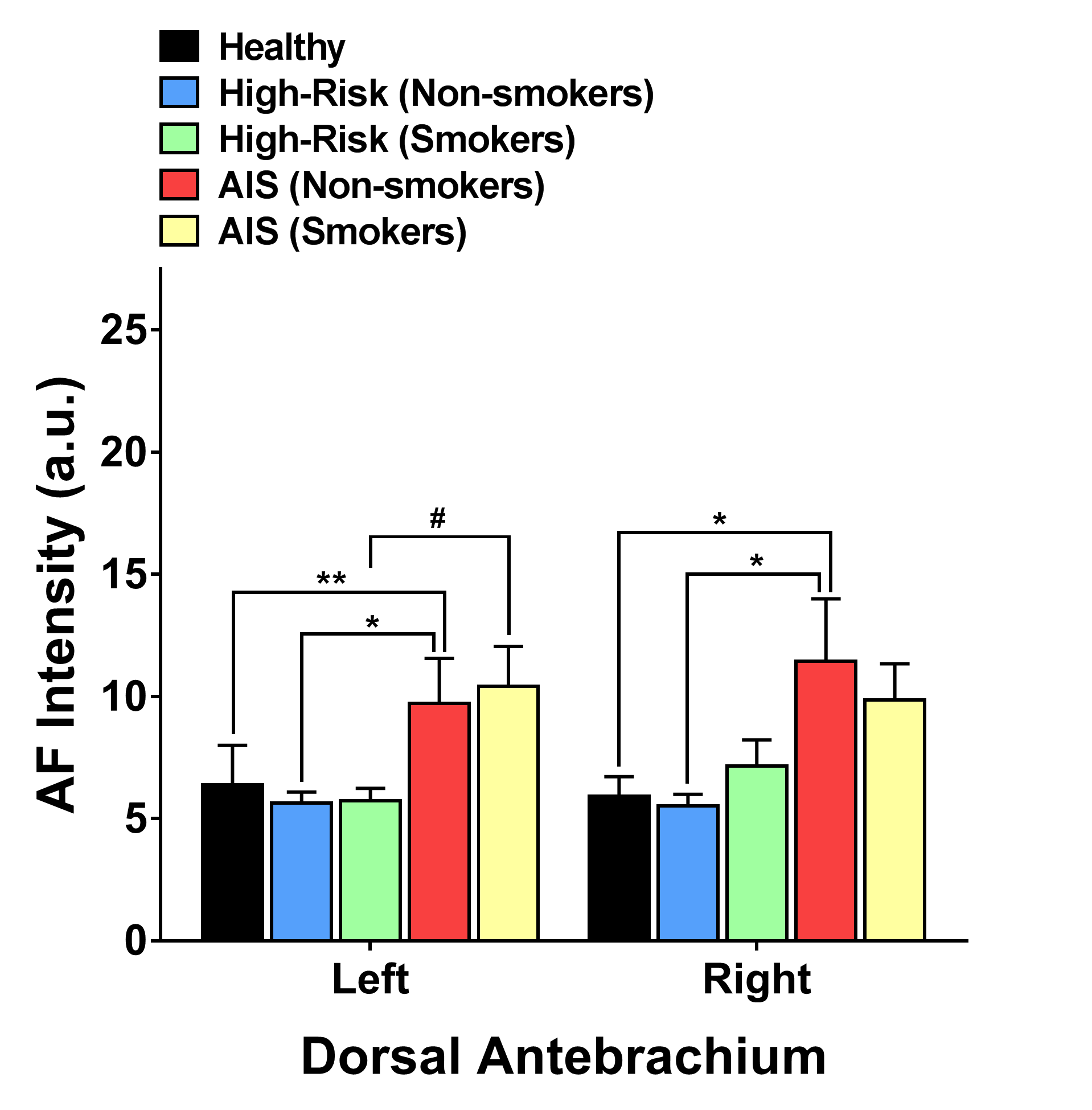

### Supplemental Fig. 3D

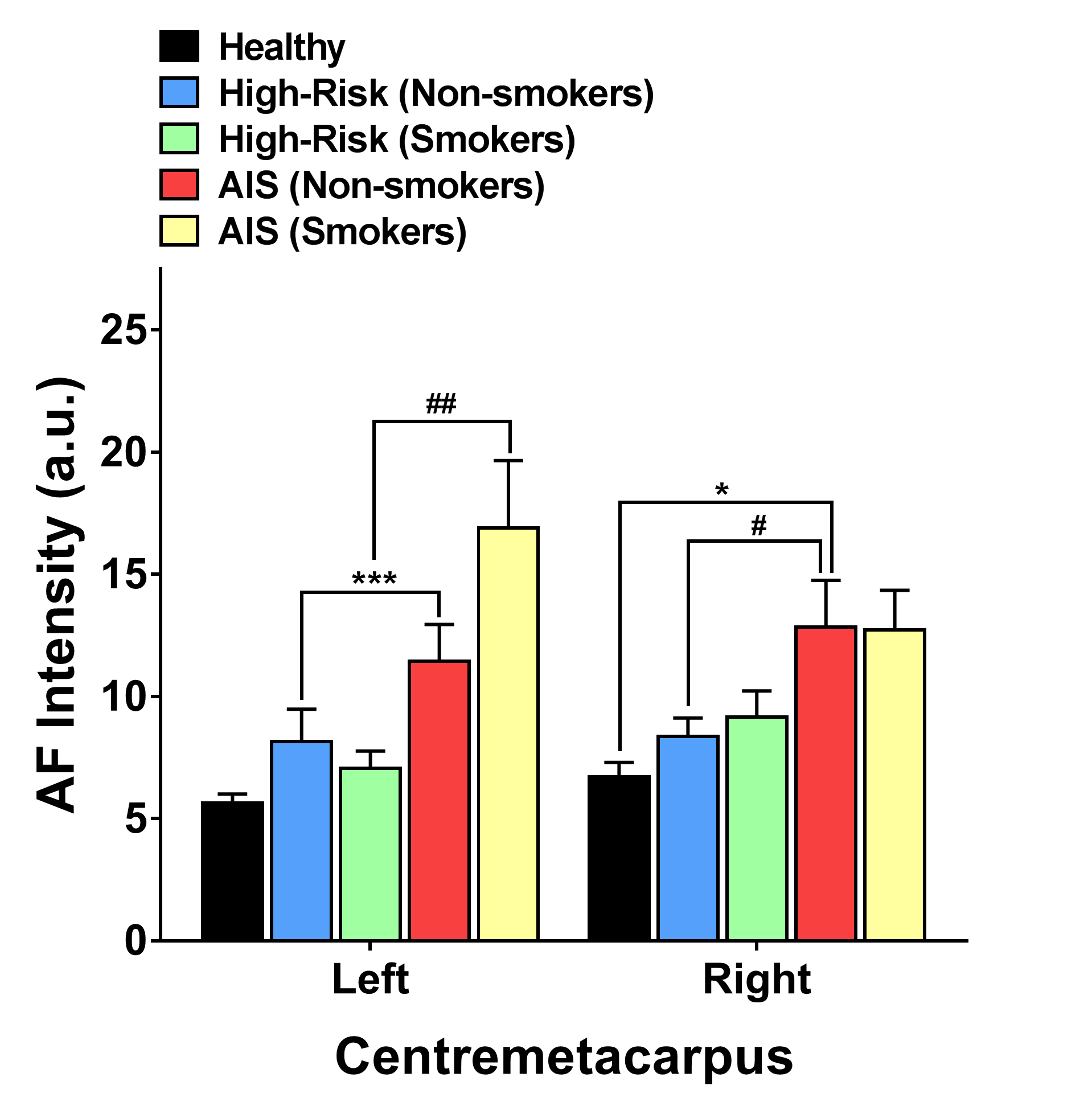

### Supplemental Fig. 3E

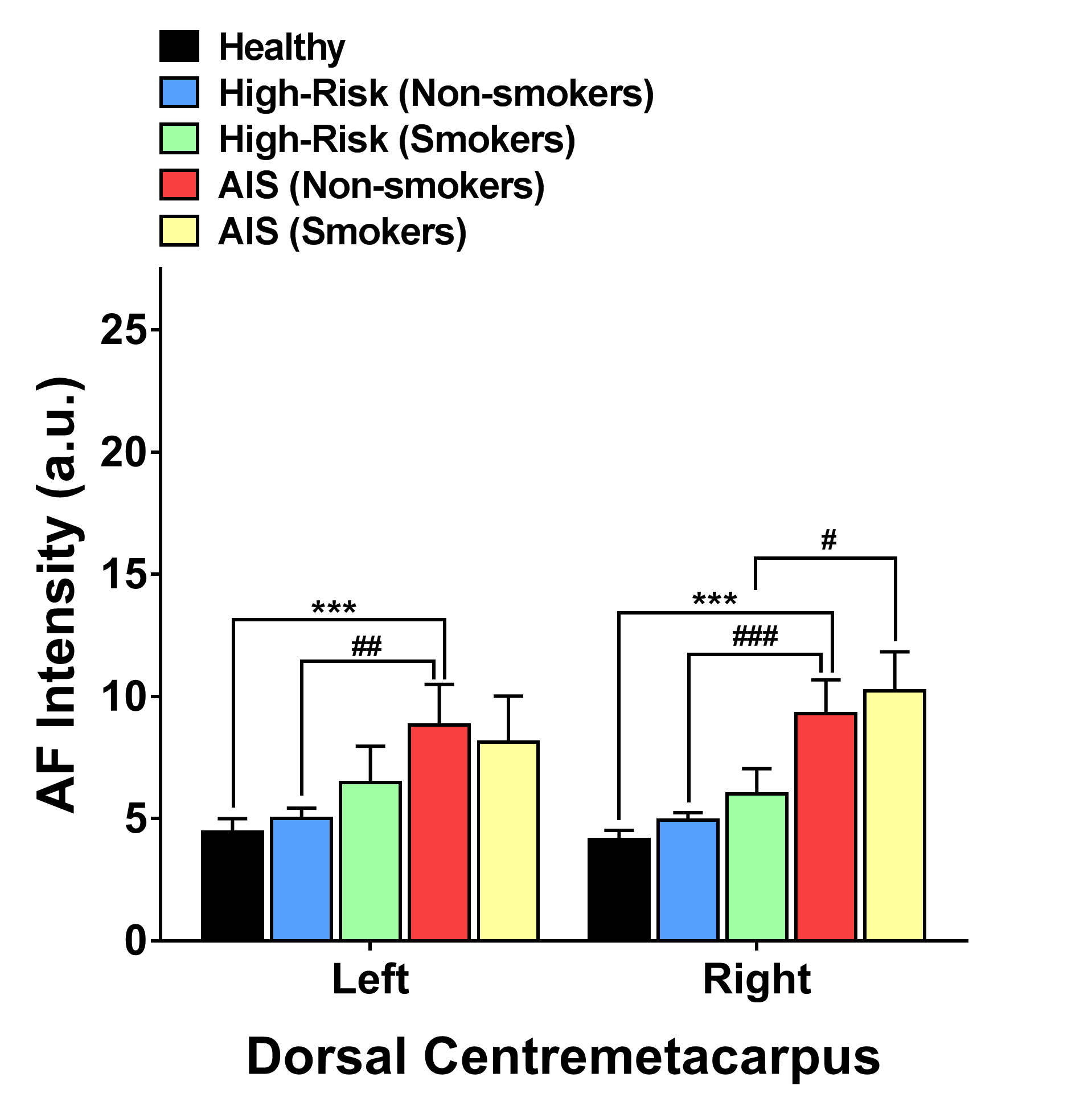

### Supplemental Fig. 4A

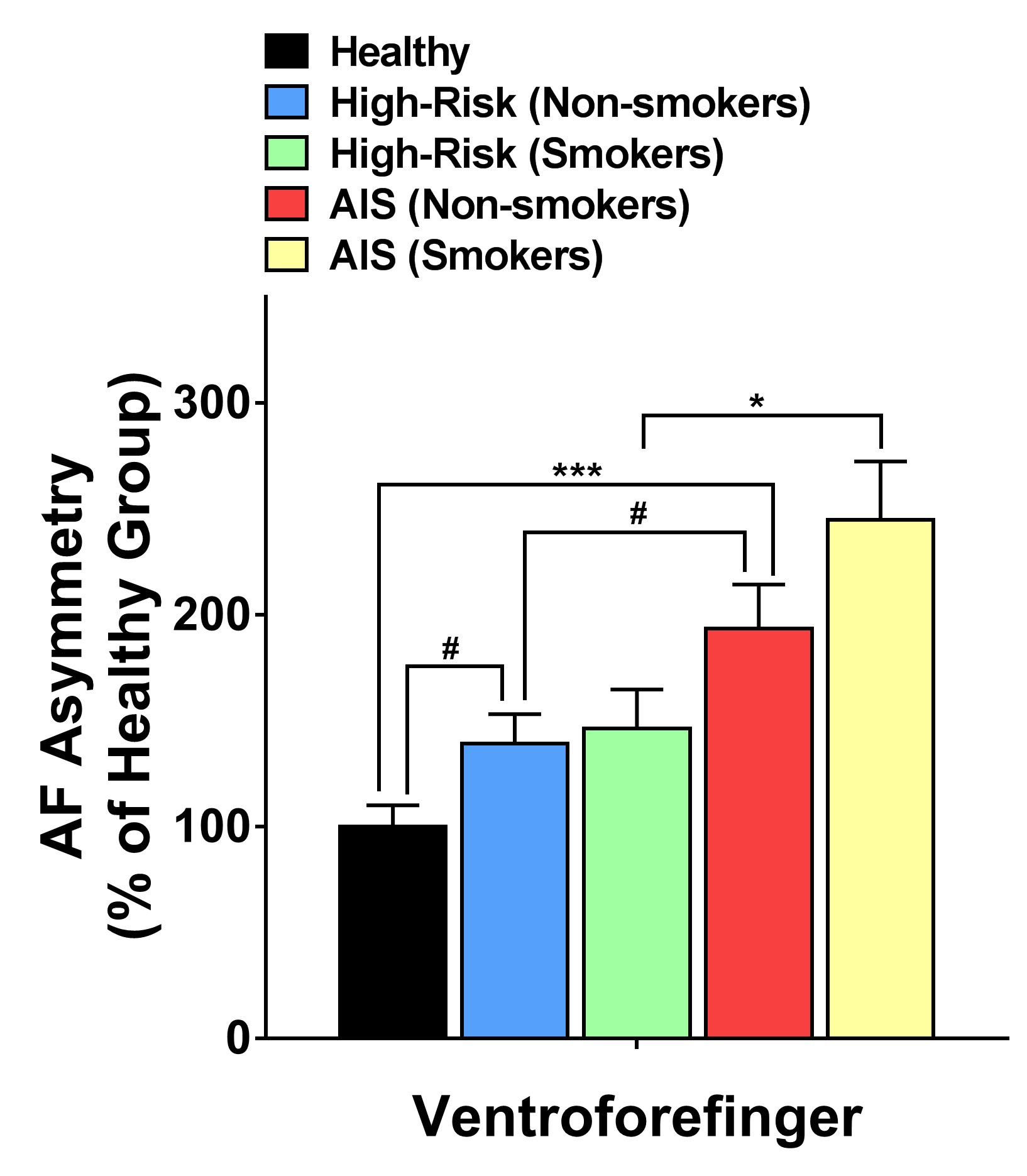

### Supplemental Fig. 4B

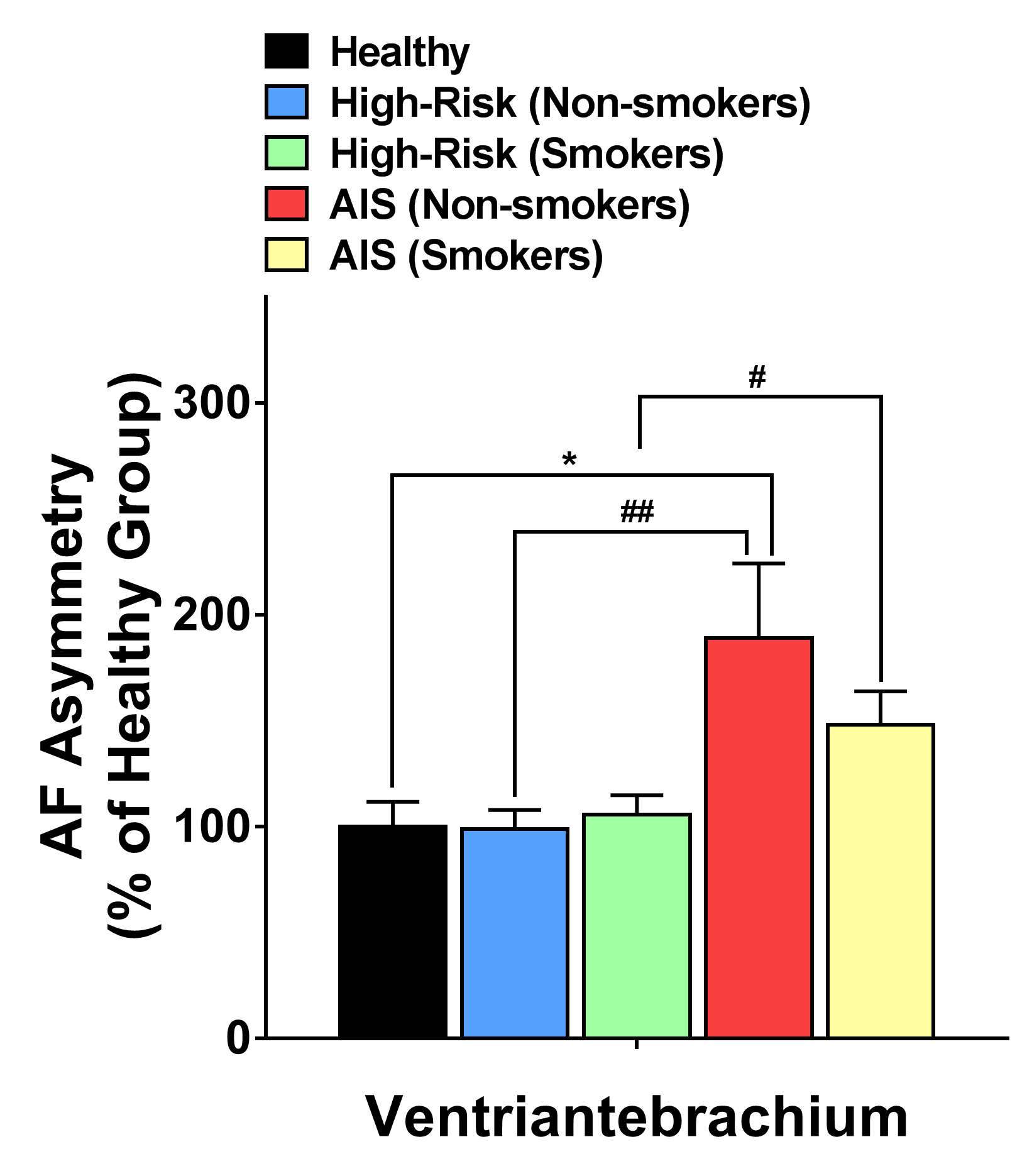

### Supplemental Fig. 4C

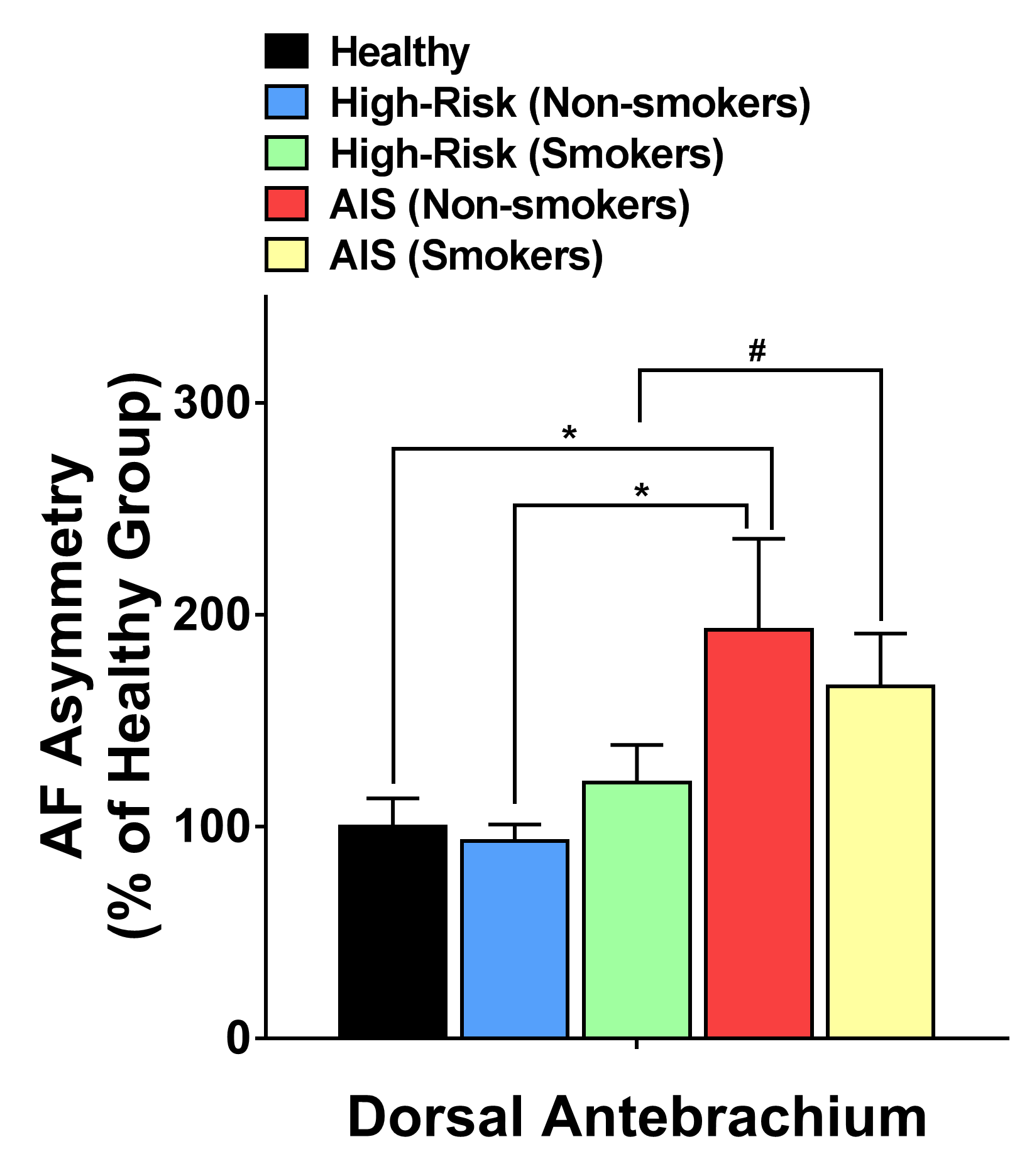

### Supplemental Fig. 4D

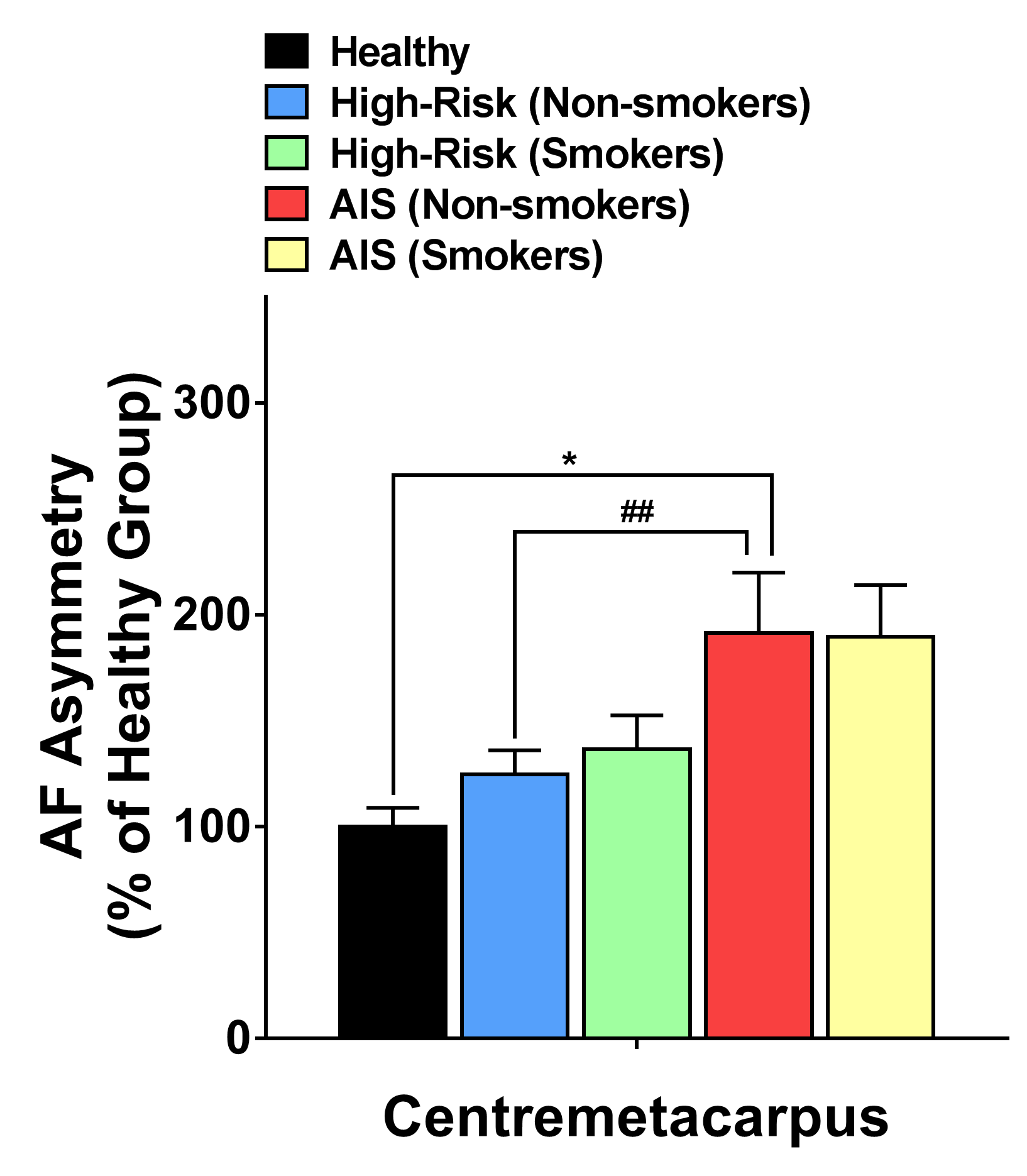

### Supplemental Fig. 4E

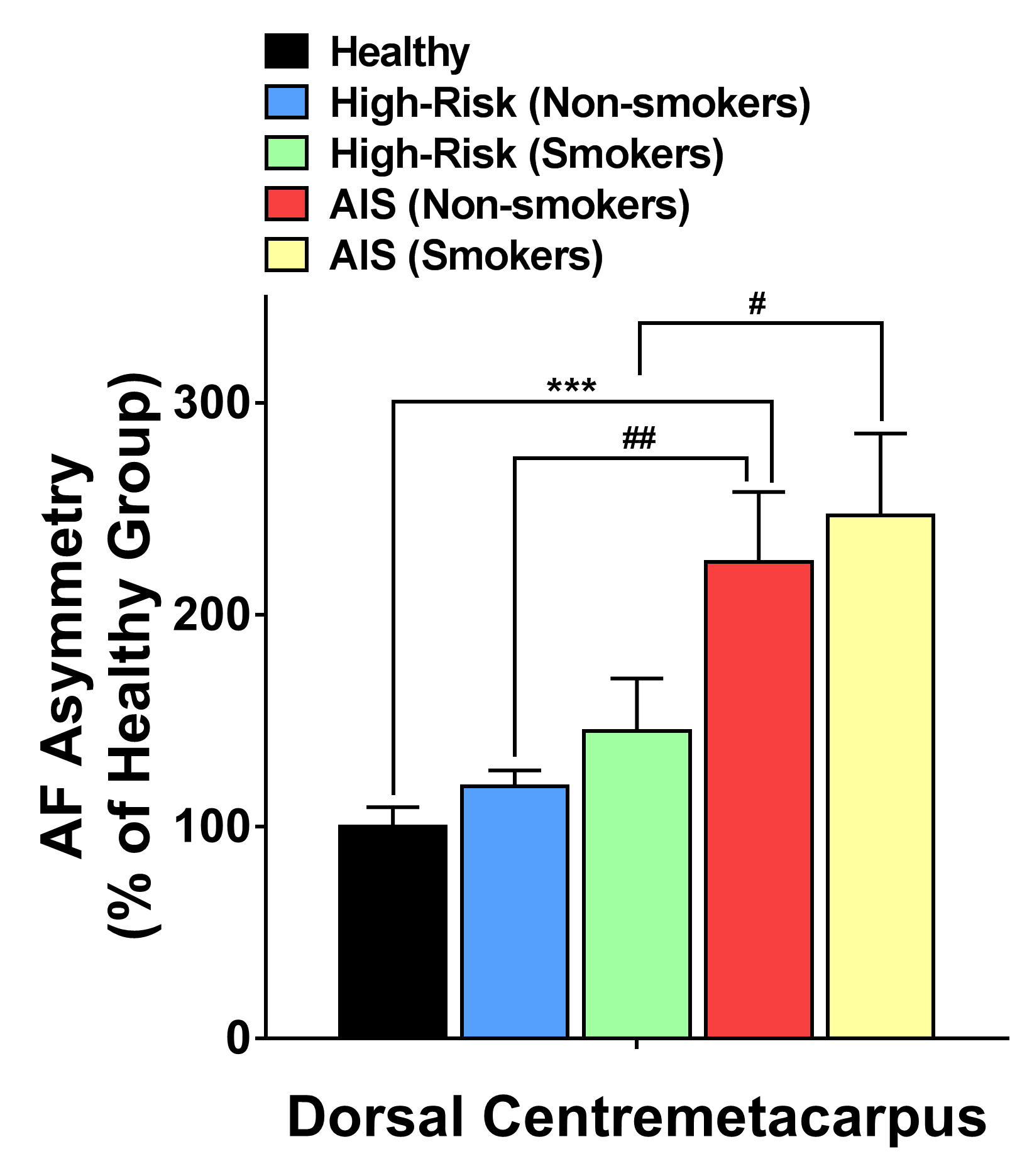
